# Circulating Lipids as Biomarkers for Diagnosis of Tuberculosis: A Multi-cohort, Multi-omics Data Integration Analysis

**DOI:** 10.1101/2024.08.06.24311536

**Authors:** Nguyen Tran Nam Tien, Nguyen Thi Hai Yen, Nguyen Ky Phat, Nguyen Ky Anh, Nguyen Quang Thu, Vu Dinh Hoa, Cho Eunsu, Ho-Sook Kim, Duc Ninh Nguyen, Dong Hyun Kim, Jee Youn Oh, Nguyen Phuoc Long

**Author notes:** These authors contributed equally to this work (Nguyen Tran Nam Tien, Nguyen Thi Hai Yen). Corresponding authors: Nguyen Phuoc Long.

## Abstract

**Background:** Circulating immunometabolic biomarkers show promise for the diagnosis and treatment monitoring of tuberculosis (TB). However, biomarkers that can distinguish TB from nontuberculous mycobacteria (NTM) infections, latent tuberculosis infection (LTBI), and other lung diseases (ODx) have not been elucidated. This study utilized a multi-cohort, multi-omics approach combined with predictive modeling to identify, validate, and prioritize biomarkers for the diagnosis of active TB.

**Methods:** Functional omics data were collected from two discovery cohorts (76 patients in the TB-NTM cohort and 72 patients in the TB-LTBI-ODx cohort) and one validation cohort (68 TB patients and 30 LTBI patients). An integrative multi-omics analysis was performed to identify the plasma multi-ome biosignatures. Machine learning-based predictive modeling was then applied to assess the performance of these biosignatures and prioritize the most promising candidates.

**Results:** Conventional statistical analyses of immune profiling and metabolomics indicated minor differences between active TB and non-TB groups, whereas the lipidome showed significant alteration. Muti-omics integrative analysis identified three multi-ome biosignatures that could distinguish active TB from non-TB with promising performance, achieving area under the ROC curve (AUC) values of 0.7–0.9 across groups in both the discovery and validation cohorts. The lipid PC(14:0_22:6) emerged as the most important predictor for differentiating active TB from non-TB controls, consistently presenting at lower levels in the active TB group compared with counterparts. Further validation using two independent external datasets demonstrated AUCs of 0.77–1.00, confirming the biomarkers’ efficacy in distinguishing TB from other non-TB groups.

**Conclusion:** Our integrative multi-omics reveals significant immunometabolic alteration in TB. Predictive modeling suggests lipids as promising biomarkers for TB-NTM differential diagnosis and TB-LTBI-ODx diagnosis. External validation further indicates PC(14:0_22:6) as a potential diagnostic marker candidate for TB.

**Summary:** Our multi-cohort, multi-omics data integration and predictive modeling identified reliable biomarkers and highlighted the importance of circulating lipids for distinguishing tuberculosis (TB) from complex conditions with similar clinical manifestations, latent infections, and healthy individuals.

## INTRODUCTION

*Mycobacterium tuberculosis* (*Mtb*) infection, which causes TB, continues to be a critical global health burden; 7.5 million new cases were reported in 2022 [1]. Individuals with latent TB infection (LTBI), characterized by a persistent immunological response against *Mtb* without clinical symptoms, have a 5–10% lifetime risk of progression to active TB [2]. However, limited methods are available to distinguish between TB and LTBI [3, 4]. Additionally, infections caused by nontuberculous mycobacteria (NTM), which include other bacterial species of the genus *Mycobacterium*, have been increasing globally at a rate of approximately 4.1% per year [5, 6]. TB, NTM infections, and also other lung diseases (ODx) share significant overlapping clinical manifestations and imaging features [7, 8], complicating the accurate diagnosis of TB. This complexity contributes to an estimated 3 million cases being missed annually [1, 9].

Sputum culture is recommended for the microbiological confirmation of TB or NTM infections [4, 10-13]. However, this method can take 1–2 months to yield results and is less effective for patients unable to consistently produce sufficient sputum samples [4, 14]. Consequently, the World Health Organization (WHO) has recommended the development of non-sputum-based diagnostic tests with high sensitivity and specificity, utilizing more easily obtainable samples such as breath condensate, urine, and blood [15]. The QuantiFERON-TB Gold In-Tube assay can differentiate TB from NTM with moderate accuracy [16, 17], but it is not effective for distinguishing TB from LTBI [3], underscoring the importance of clinical manifestations in clinical decision-making. Additionally, medical imaging is insufficient for differential diagnosis among TB, NTM, and ODx [8, 18]. A universal diagnostic test capable of differentiating TB from NTM, other lung diseases, LTBI, and healthy individuals remains necessary [14]. Circulating immune and metabolic biomarkers hold promise for this purpose [19-23]. For example, Hur *et al.* identified several cytokines, such as interleukin-2, as potential adjunctive biomarkers for distinguishing TB from NTM [19]. In a previous study, we demonstrated that phosphatidylcholines (PC) and ether derivatives [PC(O-)] are potential biomarkers for differentiating TB patients from individuals with LTBI and healthy controls [21]. Notably, lipid-related genes performed similarly compared to well-established gene expression biomarkers [24]. However, previous studies on immunometabolic biomarkers have often been exploratory and generally lack external validation. Further studies are needed to address these gaps, enhance our understanding of the metabolic alterations associated with host immune responses in these infections, and support the development of novel diagnostic tools.

In this study, we utilized an integrative multi-omics analysis combined with machine learning (ML)-based predictive modeling, in a multi-cohort design to identify, prioritize, and validate potential biomarkers for differentiating TB from NTM, LTBI, and ODx. A supervised multi-omics integration method was implemented to identify highly correlated and discriminant biomarkers, forming multi-ome biosignatures. These biosignatures were thoroughly validated using the ML models. Additionally, we externally validated the key biomarkers across multiple public data cohorts. The application of multi-omics investigation provided more profound insights into the metabolic and immune responses associated with TB and NTM infections. Overall, our comprehensive approach lays the groundwork for the use of non-sputum-based biomarkers in the differential diagnosis of TB.

## METHODS

### Patient recruitments

This study is part of a prospective multi-center TB cohort named the “Center for Personalized Precision Medicine of Tuberculosis (cPMTb)”, initiated in 2018 [25]. The Institutional Review Board of Guro Hospital, Korea University (No. 2017GR0012) evaluated and approved the protocol for gathering patients’ medical information and biospecimens for the analysis. This study included two discovery cohorts and one validation cohort, with plasma samples and medical information obtained from the Biobank of Korea University Guro Hospital. The study was conducted in accordance with the Declaration of Helsinki, and all participants provided written informed consent.

Potentially eligible patients were individuals who visited the outpatient clinic and had suspected TB based on clinical assessment, X-ray, or computed tomography. They underwent additional evaluations to reach final diagnoses of TB, NTM, or ODx. TB was diagnosed in accordance with the WHO guidelines for drug-susceptible TB, considering clinical symptoms, sputum smear microscopy and culture tests, radiological examination, and the GeneXpert MTB/RIF assay. The diagnosis of NTM was conducted according to the Clinical Practice Guideline of the American Thoracic Society, European Respiratory Society, European Society of Clinical Microbiology and Infectious Diseases, and Infectious Diseases Society of America [10]. Participants with positive results in the Interferon Gamma Release Assay but without clinical or radiographic evidence of active TB were diagnosed with LTBI. The ODx group included patients initially suspected to exhibit active TB but ultimately diagnosed with other conditions, such as lung cancer or pneumonia. Individuals receiving immunosuppressive therapy and those with severe chronic liver or renal diseases were excluded. TB and NTM patients, as well as LTBI individuals with malignant diseases, were also excluded.

Unstimulated plasma cytokine profiling, plasma metabolomics and lipidomics data were acquired for the two discovery cohorts. Prospectively collected samples available from the biobank were subsequently used for validation. The plasma metabolomics and lipidomics data were acquired using the same approach in the validation cohort. Detailed sample preparation, data acquisition, data preprocessing and data treatment are described in the Supplementary Material.

### Exploratory data analysis

We first used the principal component analysis (PCA) to illustrate the different clusters of samples from groups. Herein, MS-derived data were subjected to median normalization, log10- transformation, and Pareto scale, and MFI data was log10-transformed and Pareto scaled. For differential abundance analyses, the normalized and log10-transformed metabolomics, lipidomics, and log10-transformed immune profiling were submitted to a linear regression model with age- and body mass index (BMI)-adjustment using the *limma* method. Differential analytes of cytokines/chemokines/growth factors (immune factors), metabolites, and lipids were determined based on a P-value cut-off of 0.05 and a false discovery rate (FDR) cut-off of 0.25. These data analyses were performed in MetaboAnalyst 6.0 [26]. The heatmap of differential analytes was plotted using the *ComplexHeatmap* package version 2.14.0 [27].

### Multi-omics integration

We used supervised integrative multi-omics methods, i.e., the data integration analysis for biomarker discovery using the latent components (DIABLO) methods [28]. The analysis was implemented via the *mixOmics* package version 6.25.1 in R 4.2.3, respectively. Data normalization according to three omics data layers was performed using the same approach as differential analysis. The platform intrinsically executed data scaling. The distance method and the number of components were selected based on a minimal (balanced) error rate using a 10-fold cross-validation (CV) repeated 20 times. The circos plot representing the average levels between active TB and non-TB groups and correlations between selected biomarkers in the first component was used. The heatmap of component 1 selected biomarkers was also visualized via the *ComplexHeatmap* package version 2.14.0 [27]. The area under the receiver operating characteristic curve (AUC) of component 1 selected biosignatures of three data blocks, and selected biosignatures of all components within each data layer were obtained via a 10-fold 20-repeated CV. The classification performance was considered acceptable, good, very good, and excellent when 0.6 ≤ AUC < 0.7, 0.7 ≤ AUC < 0.8, 0.8 ≤ AUC < 0.9, and AUC ≥ 0.9, respectively. These criteria were also applied to other parts related to the classification between active TB and non-TB groups below.

### Biosignatures predictive performance evaluation

To assess cross-cohort predictability, we retrieved the component 1 DIABLO-selected multi-ome biosignature in one of three comparisons and assessed their potential to differentiate between groups in the two remaining scenarios. We also intersected the three multi-ome biosignatures to form a consensus biosignature consisting of shared biomarkers in at least 2/3 multi-ome biosignatures. The validation cohort was then used to evaluate the predictability of three multi-ome biosignatures and the consensus biosignatures. Besides, the clinical covariates (age, gender, and BMI) were also included in the input data. Three ML models of partial least squares discriminant analysis (PLS-DA), support vector machines (SVM), and random forest (RF) were implemented. Data treatment and normalization were in line with the multi-omics analysis. We split the data into a training and a testing set with a ratio of 7:3. The standard scaler method was used in the SVM and PLS-DA models to scale the continuous variables, separately in the training and testing sets. The ML models were developed and validated with 5-fold CV repeated 5 times using *caret* (version 6.0-94) and *pROC* (version 1.18.5) packages in R 4.2.3 [29, 30]. For each case, we also calculated the variable importance score.

### Biomarker external validation

We validated the most potential biomarkers by using data from the validation cohort; public data from our previous work on untargeted lipidomics-based biomarkers discovery for TB diagnosis (Long *et al.*) [21], and a targeted metabolomics study for TB diagnosis biomarker identification by Cho *et al.* [31]. The potential biomarkers were evaluated based on the AUC through the classical univariate receiver operating characteristic (ROC) curve analyses in MetaboAnalyst 6.0 [26]. We also used a one-point-based quantification method to acquire the concentration of the most potential candidates from all samples across our three cohorts. Using concentration data, the univariate ROC analysis and logistic regression (repeated 5 times, with or without adding age, gender, and BMI) were conducted in MetaboAnalyst 6.0 [26] and *caret* package (version 6.0-94) coupled with *pROC* (version 1.18.5) package in R 4.2.3, respectively [29, 30]. The relative abundance/concentration of the biomarkers across groups in our three cohorts and public studies was also examined.

## RESULTS

### Baseline characteristics

Our study comprised two discovery cohorts (TB-NTM and TB-LTBI-ODx cohorts) and one validation cohort (TB-LTBI cohort). The study design and workflow are shown in **Figure 1**, and the clinical characteristics of the two discovery cohorts are summarized in **Table 1**.

**Figure 1.**
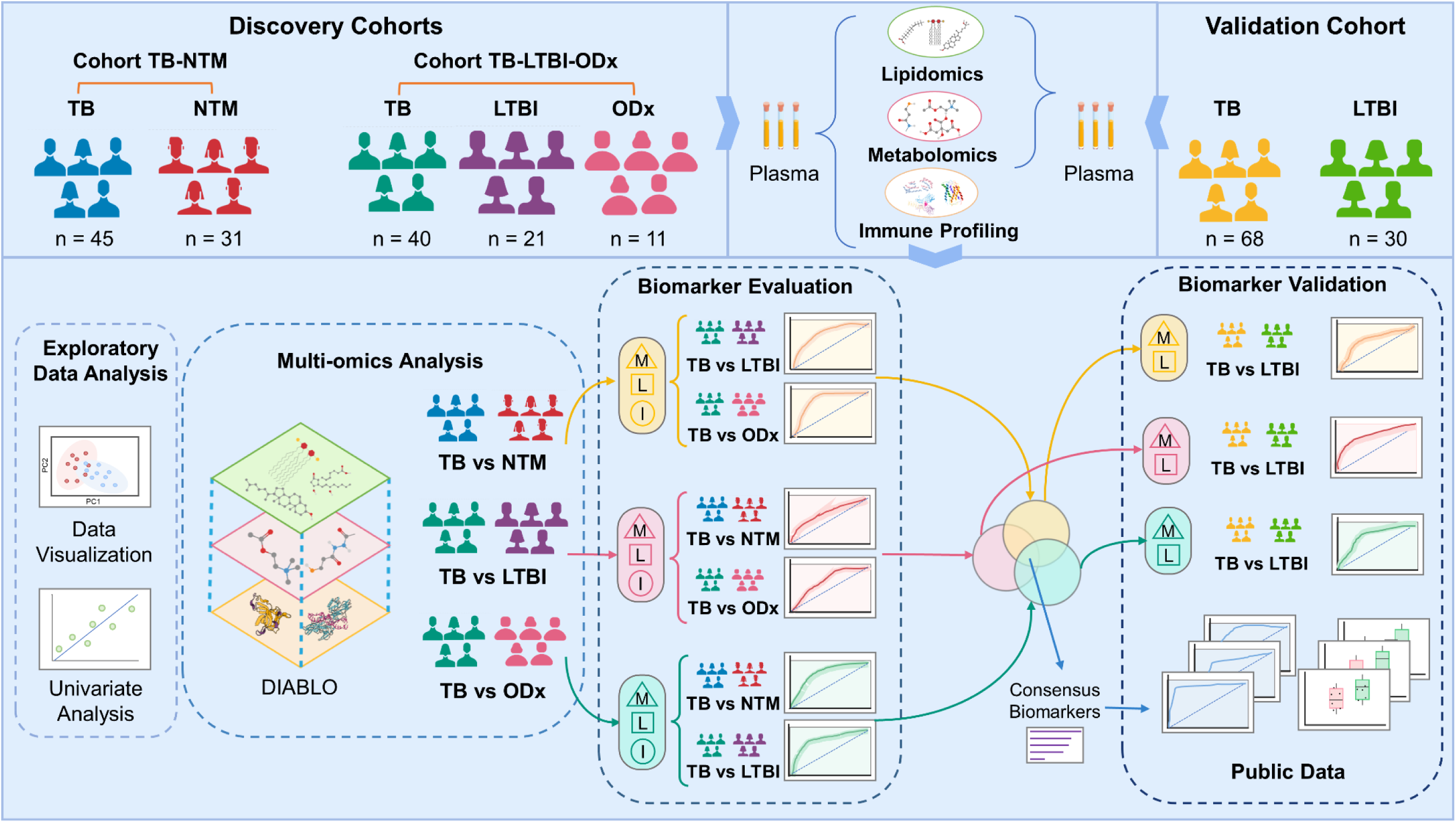
Study design and workflow. Abbreviations: TB: tuberculosis; NTM: nontuberculous mycobacteria infection; LTBI: latent tuberculosis infection; and ODx: other lung diseases.

**Table 1.**
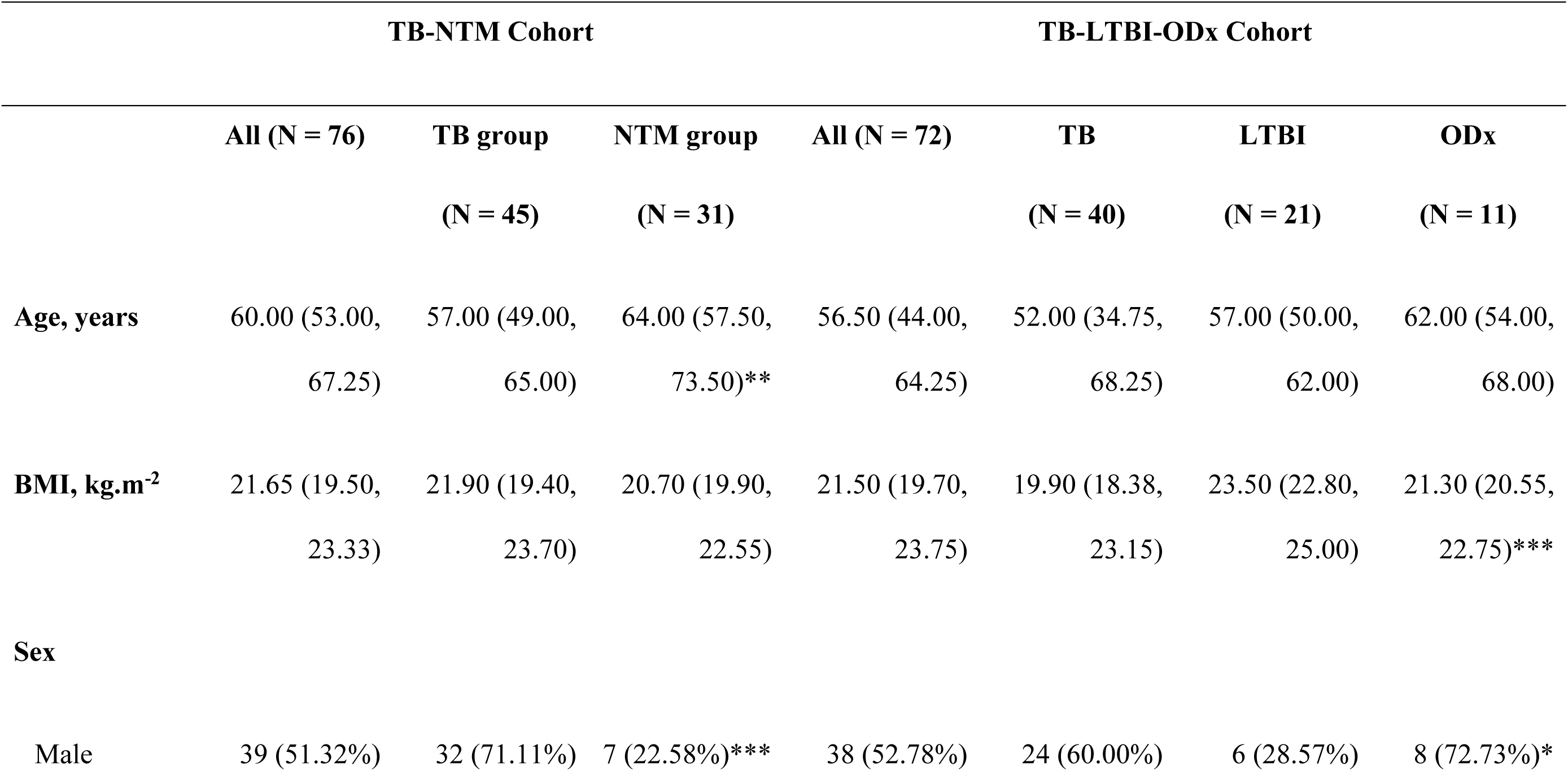
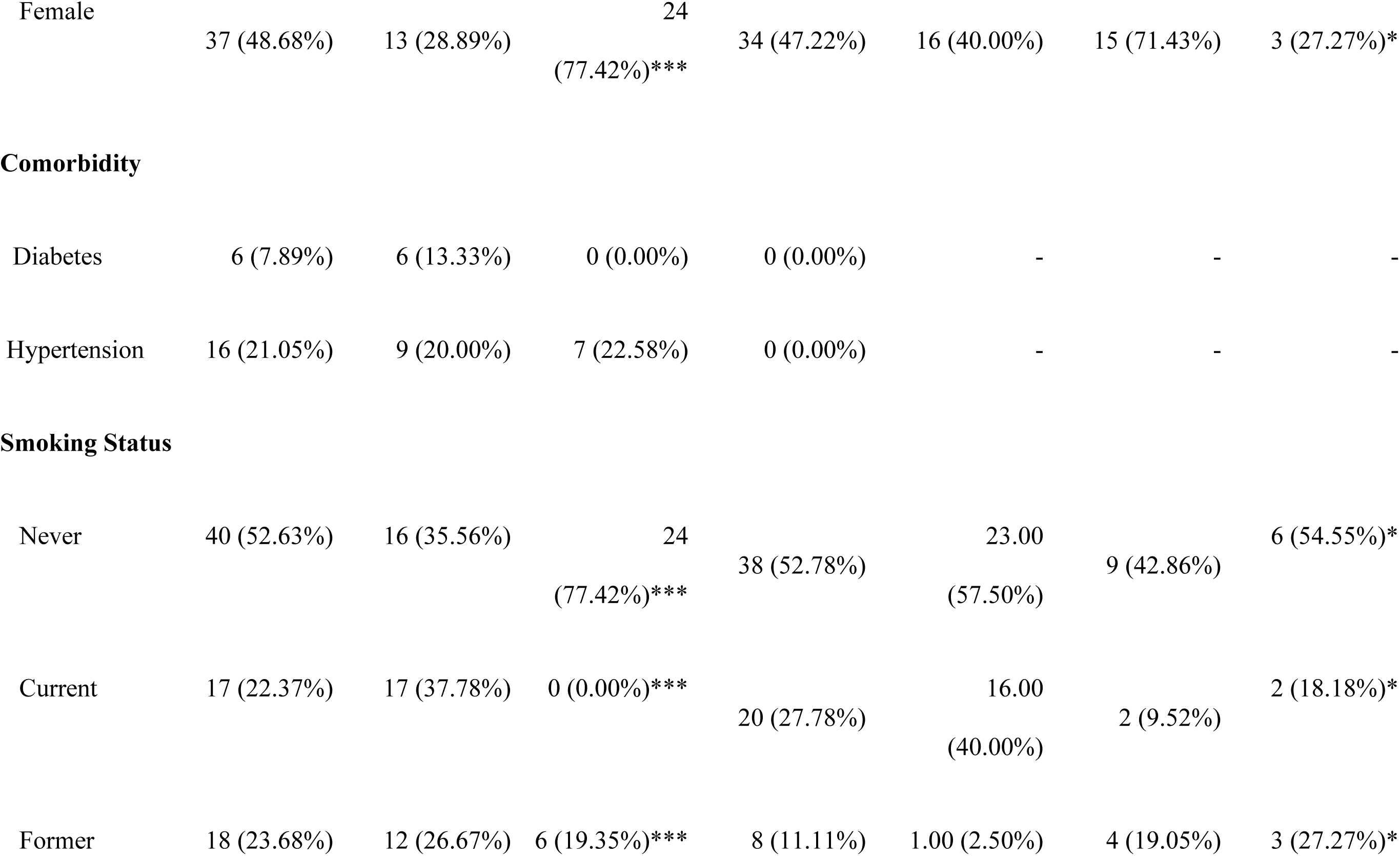
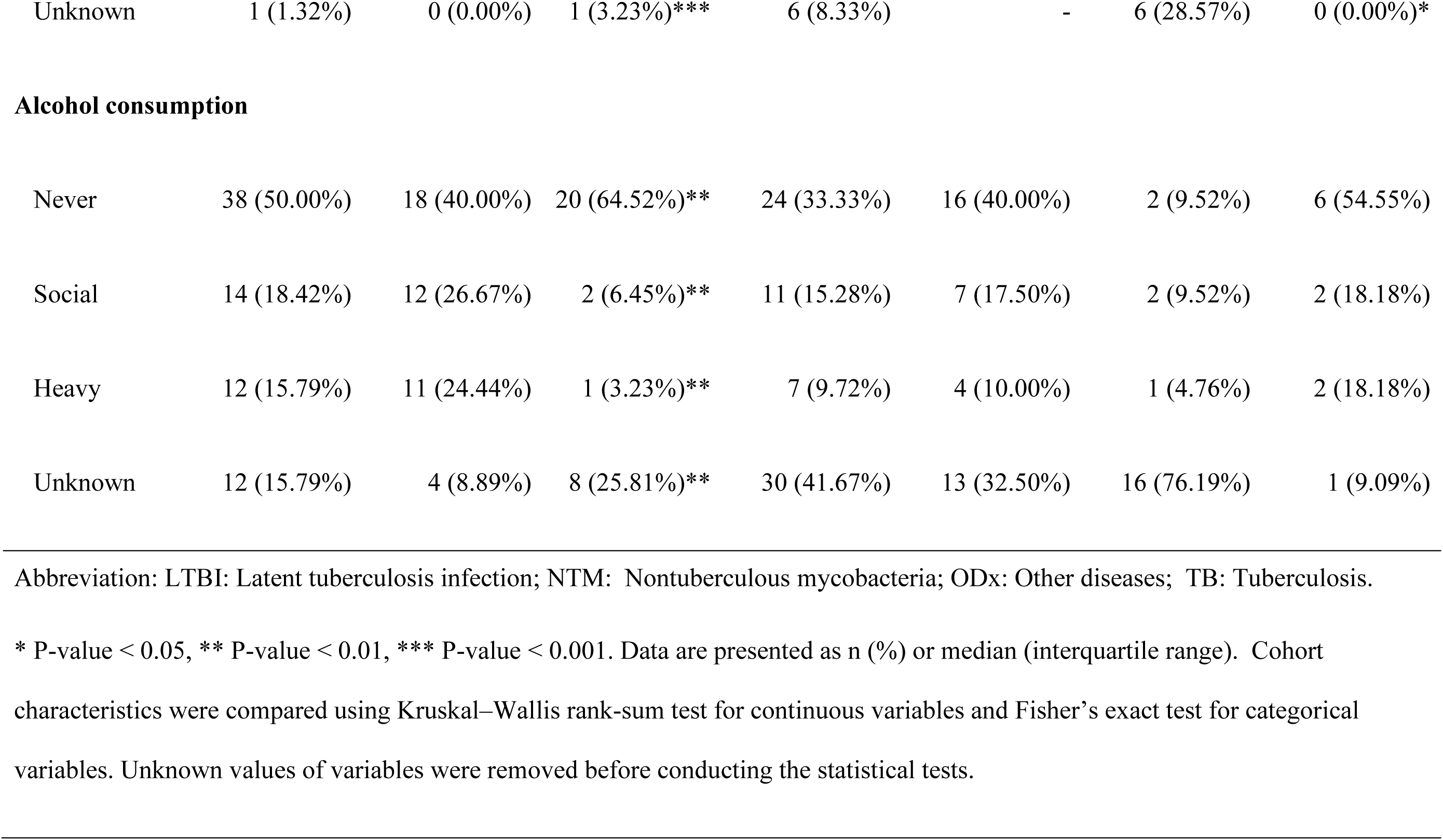
Clinical characteristics of the two discovery cohorts.

The TB-NTM cohort included 45 pulmonary TB patients and 31 NTM patients. The age of TB patients was significantly lower than that of individuals with NTM. The TB group also had a higher proportion of men compared with the NTM group. Six NTM patients and no TB patients had diabetes. More individuals in the TB group smoked and consumed alcohol than those in the NTM group. There were no significant differences between the two groups in terms of BMI and the proportion of individuals with hypertension. The TB-LTBI-ODx cohort comprised 40 patients with pulmonary TB, 21 patients with LTBI, and 11 patients with ODx, including two lung cancer patients. There were no significant age differences among the groups in this cohort. Patients with LTBI had the highest BMI, followed by those with ODx and TB. The ODx group had the highest proportion of men, whereas the LTBI group had the highest proportion of women. None of the patients in the TB-LTBI-ODx cohort had hypertension or diabetes. TB patients smoked more than ODx and LTBI patients, but there were no significant differences in alcohol consumption among the participants in this cohort.

The validation cohort consisted of 68 TB patients and 30 LTBI participants, with detailed characteristics presented in **Supplementary Table S1**. The two groups exhibited significant differences in all characteristics (i.e., BMI, sex ratio, hypertension and diabetes prevalence, smoking status, and alcohol consumption), except for age. This variability made the cohort suitable for validation purposes.

### Immune profiles, metabolome, and lipidome between active TB and non-TB groups

PCA was performed to explore sample tendencies regardless of origin, using immune profiling, metabolomics, and lipidomics data. **Supplementary Figure S1** shows all the scores plots of the two cohorts. In the TB-NTM cohort, no significant differences were observed between the two groups based on immune and metabolome profiles, although a slight difference in lipidome profiles was noted between the TB and NTM groups. In the TB-LTBI-ODx cohort, PCA score plots revealed a subtle separation between TB and LTBI samples across the three data modalities. For the TB and ODx comparison, immune profiles demonstrated greater separation compared with the separation of metabolome and lipidome profiles.

Next, univariate analysis was performed using a linear model with clinical covariate adjustment to identify differential analytes. In the TB-NTM cohort, six immune factors were significantly up-regulated in TB, including pro-inflammatory interleukins (IL-6, IL-8, IL-15, IL-18), CXC chemokine of GRO alpha, and VEGF-A (**Figure 2A**). Additionally, 14 metabolites showed significantly different abundances between the TB and NTM groups. Five lysophosphatidylcholines (LPC), citrate, hippurate, and acetylserine were down-regulated, while indolelactate was up-regulated in the TB group. Notably, 134 lipids from the lipidome analysis were statistically different between TB and NTM patients, with changes ranging from 1.2- to 2-fold. The alterations in the lipidome were primarily due to the consistently higher levels of triacylglycerols (TG) and lower levels of sphingomyelins (SM), PC, PC(O-), and ether-linked phosphatidylethanolamines [PE(O-)] in the TB group.

**Figure 2.**
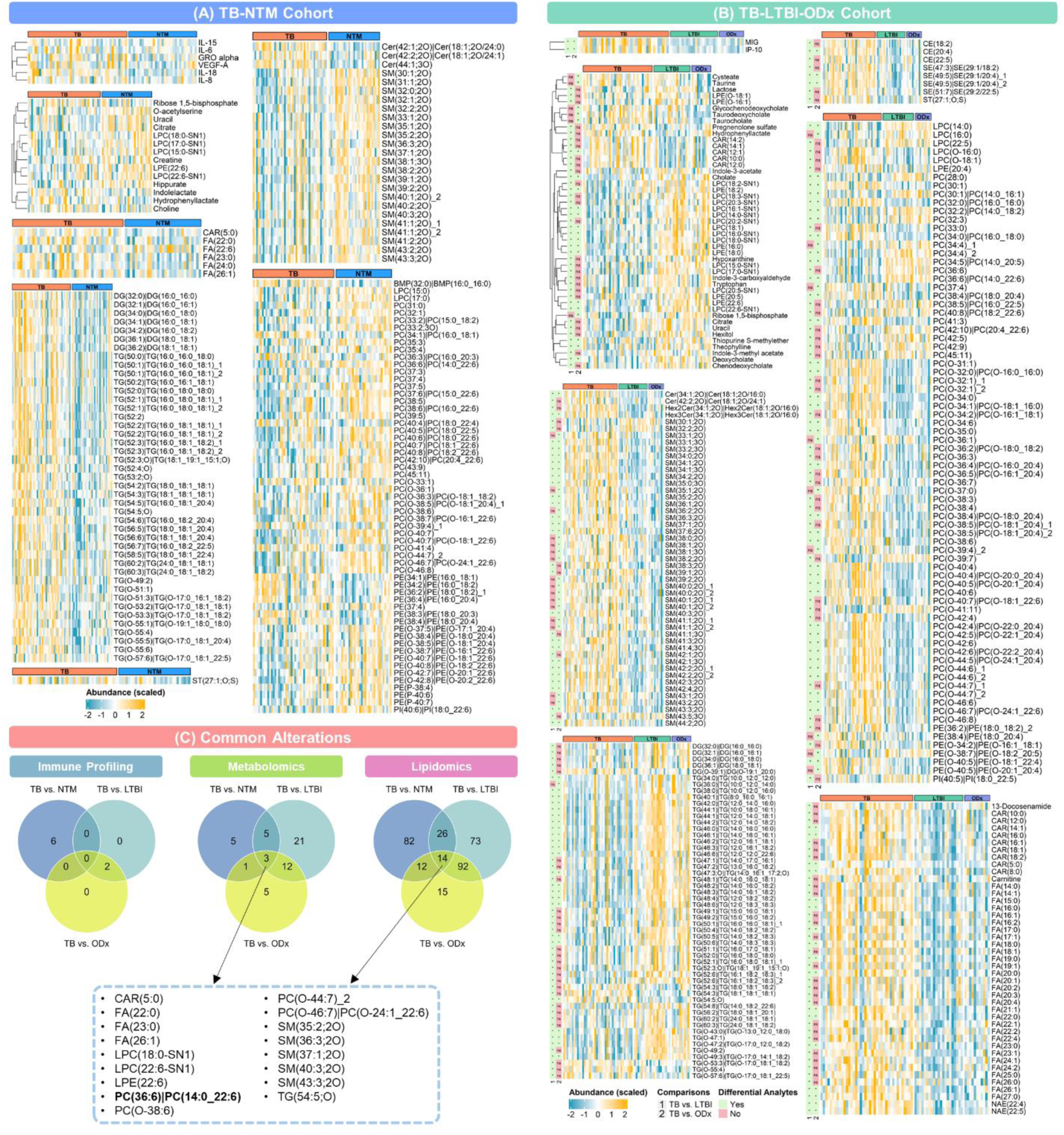
Heatmap of differential analytes in the discovery cohorts. **(A)** The TB-NTM cohort. **(B)** The TB-LTBI-ODx cohort. **(C)** Common differential analytes between 2 cohorts. Abbreviations: TB: tuberculosis; NTM: nontuberculous mycobacteria infection; LTBI: latent tuberculosis infection; and ODx: other lung diseases.

In the TB-LTBI-ODx cohort, the adjusted linear model using immune profiling data found only two significantly up-regulated immune factors, MIG and IP-10, when comparing TB and LTBI participants (**Figure 2B**). Meanwhile, 41 differential metabolites were found in the metabolomics analysis. Increased abundances of five acylcarnitines and decreased abundances of LPC and LPE [e.g., LPC(14:0-SN1), LPE(16:0), LPC(22:6-SN1), LPE(22:6)] were observed in the TB group. Additionally, the lower levels of two primary bile acids (cholate and chenodeoxycholate), three metabolites related to tryptophan metabolism (tryptophan, indole-3-acetate, and indole-3-methyl acetate), hypoxanthine, and uracil were also detected in the TB patients. Lipidomics analysis yielded 205 (137 up- and 68 down-regulated levels) differential lipids. For the comparison between TB and LTBI, lipidome alterations in the TB group were mainly characterized by higher abundances of fatty acids (12 saturated and 18 unsaturated), acylcarnitines, cholesteryl esters, SM, and PC(O-), and lower levels of diacylglycerols (DG) and TG. Between the TB and ODx groups, two differential immune factors (IP-10 and MIG) were found with increased levels (approximately 3-fold) in the TB group (**Figure 2B**). Regarding the metabolomics analysis, 21 significantly different metabolites were found. A majority of them (10/21) belong to LPC [e.g., LPC(14:0-SN1) and LPC(22:6-SN1)] and LPE [e.g., LPE(18:0) and LPE(22:6)] subclasses with lower abundances in the TB group. Although bile acids (cholate, deoxycholate) decreased approximately 2 times, their conjugated forms (i.e., taurocholate and taurodeoxycholate, and glycochenodeoxycholate) increased 2- to 6-fold in the TB group. Levels of two amino acids (cysteate and taurine) were slightly increased. There were 133 lipid species with differential abundance between the TB and ODx groups. The same trend of alteration as in the case of TB versus LTBI was also observed, i.e., increased acylcarnitines, free fatty acids (FAs), PC(O-), SE, SM, and decreased TG in the TB group.

Among the six significant inflammation-promoting immune factors detected in the TB-NTM cohort, five cytokines were consistently up-regulated in the TB-LTBI-ODx cohort, although they did not meet the adjusted significance threshold. Common significant alterations in the metabolome and lipidome between TB and other groups included the down-regulated levels of LPC(22:6-SN1), LPE(22:6), PC(14:0_22:6); and up-regulated levels of CAR(5:0), FA(22:0), FA(23:0), FA(26:1), TG(18:1_18:1_18:1), and TG(54:5;O) (**Figure 2C**). Overall, the differences in the immune profiles and metabolomes between active TB and non-TB groups were subtle. However, the lipidome showed noticeable alterations in the active TB group compared with the non-TB control groups.

### Multi-ome biosignatures for differentiation between active TB and non-TB

Exploratory data analysis indicated that relying on a single omics approach might not be sufficient to identify a biosignature capable of effectively distinguishing active TB from non-TB controls. Therefore, we conducted an integrative multi-omics analysis using immune profiling, metabolomics, and lipidomics data. We focused on supervised multi-omics analysis to identify multi-ome biosignatures with the potential to differentiate TB from non-TB groups. The resulting multi-ome biosignatures for comparison of TB versus NTM, TB versus LTBI, and TB versus ODx were designated as the TB-NTM biosignature, TB-LTBI biosignature, and TB-ODx biosignature, respectively.

In the TB-NTM cohort, a strong correlation was observed between the lipidomics and metabolomics data layers (correlation coefficient, r = 0.78), and intermediate correlations (r = 0.48) were observed between immune profiling with metabolomics, and lipidomics data (**Supplementary Figure S2A)**. Consistent with our exploratory data analysis, a subtle separation between TB and NTM samples was revealed. Specifically, positive correlations were present between IL-18 and TGs, whereas negative correlations were observed between IL-18, IL-17e/IL-25 and PCs. The TB group exhibited a high abundance of IL-15, IL-17e/IL-25, VEGF-a, acylcarnitines, and several TGs, along with a low abundance of lipids from the PC(O-) subclass, urea, phenylacetylglutamine, and hippurate (**Figure 3A-B**). The use of two components from each data layer resulted in an AUC of 0.81 in the immune profiling block, 0.93 in the metabolomics block, and 0.95 in the lipidomics block (**Supplementary Figure S2B-D)**. Furthermore, a TB-NTM biosignature comprising 80 analytes (40 lipids, 30 metabolites, and 10 immune factors), was selected for further evaluation; it demonstrated very good performance (AUC = 0.87, P-value < 0.001) in distinguishing between TB and NTM.

**Figure 3.**
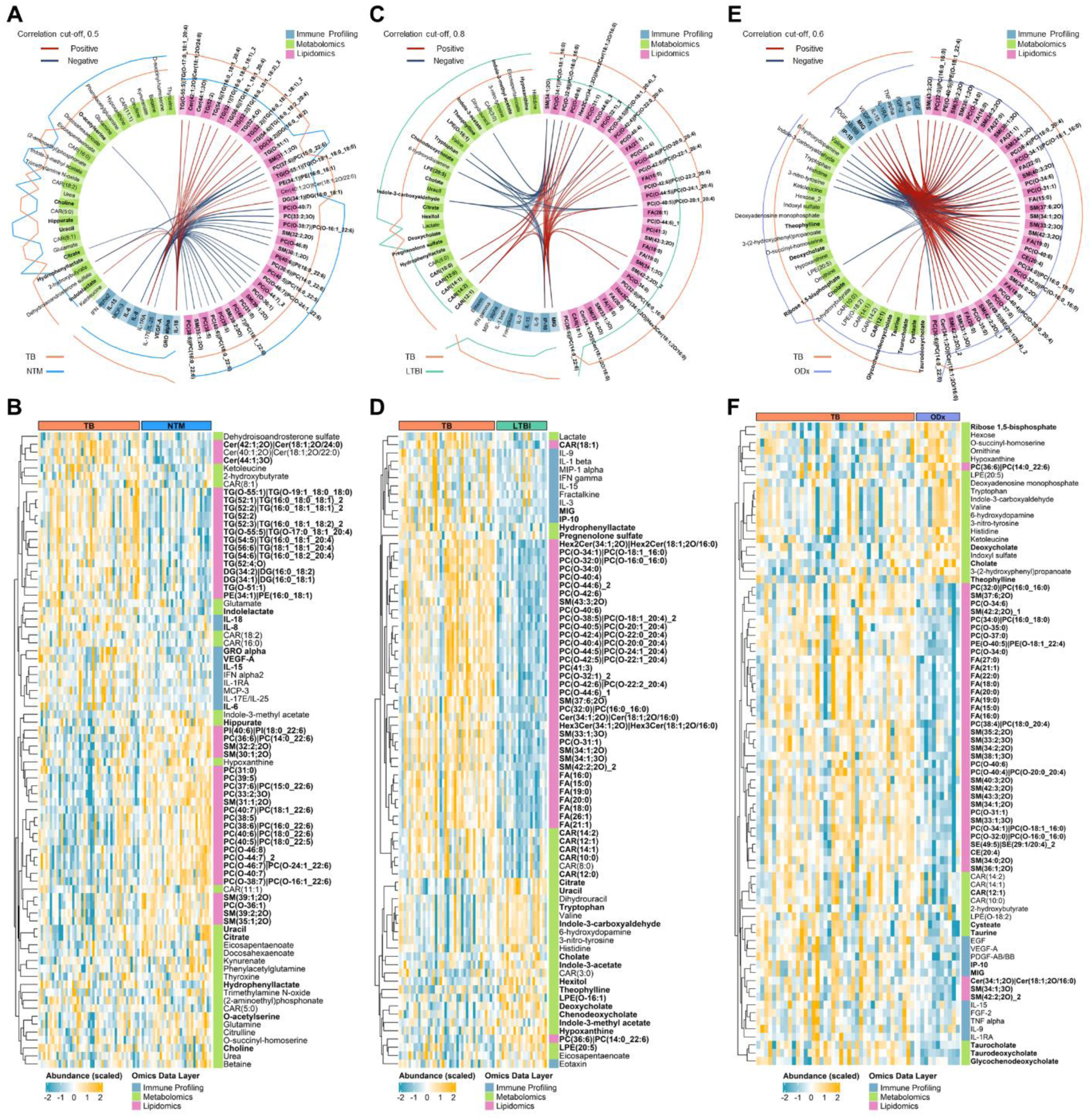
DIABLO-derived highly correlated and discriminant biosignatures for the differential diagnosis of TB-NTM, TB-LTBI, and TB-ODx. **(A)** Circos plot and **(B)** heatmap of selected biomarkers for the differential diagnosis of TB-NTM. **(C)** Circos plot and **(D)** heatmap of selected biomarkers for the differential diagnosis of TB-LTBI. **(E)** Circos plot and **(F)** heatmap of selected biomarkers for the differential diagnosis of TB-ODx. Abbreviations: TB: tuberculosis; NTM: nontuberculous mycobacteria infection; LTBI: latent tuberculosis infection; and ODx: other lung diseases.

In the TB-LTBI-ODx cohort, an integrative analysis comparing TB and LTBI revealed intermediate correlations between the different data modalities (correlation coefficient ranging from 0.57–0.63). The differentiation between the TB and LTBI groups was more distinct than that observed between the TB and NTM groups (**Supplementary Figure S3A)**. The circos plots showed the positive correlations of MIG, IL-15, and IFN-g with FAs and PC(O-)s; and negative correlations between histidine, valine, tryptophan, and deoxycholate with CAR(18:1), Cer(34:1;2O), and some PC(O-)s (**Figure 3C**). TB samples were characterized by higher levels of MIP-1a, IL-3, IL-9, FAs [e.g., FA(20:0), FA(16:0), and FA(21:1)], and some PC(O-)s [e.g., PC(O-44:5), PC(O-32:1), and PC(O-32:0)] compared with LTBI samples (**Figure 3C-D**). Meanwhile, lower levels of lauroylcarnitine, indole-3-methyl acetate, indole-3-carboxyaldehyde, uracil, and hypoxanthine were observed. Of note, the immune profiling, metabolomics, and lipidomics blocks resulted in AUCs of 0.87, 0.88, and 0.95, respectively for distinguishing TB from LTBI using two components (**Supplementary Figure S3B-D)**. The TB-LTBI biosignature was established from 37 lipids, 30 metabolites, and 10 immune factors. This biosignature demonstrated very good performance (AUC of 0.85, P-value < 0.001) for the classification of TB and LTBI.

For the comparison of TB and ODx, the lipidome and metabolome profiles exhibited a high correlation (r = 0.71), whereas the correlations of the immune profile with lipidome and with metabolome profiles were intermediate (r = 0.61 and 0.69, respectively) (**Supplementary Figure S4A**). The model was also capable of discriminating TB and ODx samples. Negative correlations of valine, histidine, 3-nitro-tyrosine, tryptophan, indole-3-carboxyaldehyde, deoxyadenosine monophosphate, and cholate with free fatty acids, several SM and PC(O-) species, IP-10, MIG, and PDGF-AB/BB was observed (**Figure 3E**). The positive correlations were determined between acylcarnitines [e.g., CAR(12:1), CAR(14:1), and CAR(14:2)] with IL-15, IL-1ra, TNF-a, PC(O-31:1), PC(O-35:0), SM(42:2;O2) and SM(38:1;3O). The abundance of selected features between the TB and ODx groups is shown in **Figure 3F**. The TB-ODx prediction model showed excellent prediction ability with lipidomics data (AUC = 0.90), while good performance was observed using the metabolomics data (AUC = 0.84) and immune profiling data (AUC = 0.72) (**Supplementary Figure S4B-D)**. The TB-ODx biosignature consisted of 80 biomarkers (40 lipids, 30 metabolites, and 10 immune factors) exhibited very good performance with an AUC of 0.86 (P-value = 0.004).

Collectively, the three multi-ome biosignatures showed considerable potential for use within circulating multi-ome profiles during TB diagnosis. We sought to establish a “consensus biosignature” by intersecting the analytes from these three biosignatures, resulting in the selection of 50 analytes. Among these analytes, IL-15, hypoxanthine, and PC(14:0_22:6) were consistently observed in all three multi-ome biosignatures (**Supplementary Figure S5**). Notably, IL-15 levels increased, whereas hypoxanthine and PC(14:0_22:6) levels declined in the TB group compared with non-TB groups. Furthermore, only the abundance of PC(14:0_22:6) showed a statistically significant difference across all three comparisons between TB and non-TB groups (**Figure 2C**).

### Independent validation showed satisfactory predictability of the multi-ome biosignatures

We assessed the predictability of the three multi-ome biosignatures in cross-cohort scenarios. Specifically, the biosignature derived from the TB-NTM cohort (TB-NTM biosignature) was independently tested to determine its ability to differentiate TB from LTBI and TB from ODx in the TB-LTBI-ODx cohort. Similarly, the TB-LTBI biosignature was evaluated for its discriminatory power to differentiate TB from NTM and TB from ODx. The TB-ODx biosignature was also tested to determine if it could distinguish between TB and NTM, as well as between TB and LTBI. Additionally, the three multi-ome biosignatures and the consensus biosignatures of metabolites and lipids were assessed for their potential to differentiate TB from LTBI in the validation cohort.

For the classification between TB and NTM, the TB-LTBI biosignature demonstrated acceptable predictability, with AUCs of 0.75, 0.78, and 0.83 in the RF, SVM, and PLS-DA models, respectively (**Table 2**, **Supplementary Figure S6A-C**). The most important features identified were PC(14:0_22:6), citrate, uracil, hydrophenyllactate, indole-3-methyl acetate, gender, and age (**Supplementary Figure S7A-C)**. Regarding the TB-ODx biosignature, the three ML models achieved AUCs of 0.80 to 0.82 in distinguishing TB from NTM patients (**Table 2**, **Supplementary Figure S6D-F**). The notable features in these models included PC(14:0_22:6), ketoleucine, PE(O-18:1_22:4), VEGF-A, FA(22:0), age, and gender (**Supplementary Figure S7D-F)**.

**Table 2.** Performance of multi-ome biosignatures and clinical covariates for the classification of active TB and non-TB.

| Biosignature | AUC ± SD | | | | | | | | |
| --- | --- | --- | --- | --- | --- | --- | --- | --- | --- |
|  | TB vs NTM Classification |  |  | TB vs LTBI Classification |  |  | TB vs ODx Classification |  |  |
|  | RF | SVM | PLS-DA | RF | SVM | PLS-DA | RF | SVM | PLS-DA |
| **TB-NTM** | | | | 0.90 ± | 0.85 ± | 0.89 ± | 0.68 ± | 0.68 ± | 0.72 ± |
| **Biosignature** |  |  |  | 0.06 | 0.03 | 0.02 | 0.16 | 0.07 | 0.06 |
| **TB-LTBI** | 0.75 ± 0.1 | 0.78 ± | 0.83 ± | | | | 0.80 ± | 0.79 ± | 0.83 ± |
| **Biosignature** |  | 0.06 | 0.05 |  |  |  | 0.12 | 0.05 | 0.07 |
| **TB-ODx** | 0.81 ± | 0.80 ± | 0.82 ± | 0.94 ± | 0.94 ± | 0.94 ± | | | |
| **Biosignature** | 0.09 | 0.04 | 0.01 | 0.04 | 0.03 | 0.04 |  |  |  |
| Abbreviation: AUC: Area under the receiver operating characteristic curve; LTBI: Latent tuberculosis infection; NTM: Nontuberculous mycobacteria; PLS-DA: Partial least squares discriminant analysis; ODx: Other diseases; TB: Tuberculosis; RF: Random forest; SD: Standard deviation; SVM: Support vector machines. |  |  |  |  |  |  |  |  |  |

For TB and LTBI discrimination, the TB-NTM biosignature demonstrated satisfactory performance, encompassing RF (AUC = 0.90), SVM (AUC = 0.85), and PLS-DA (AUC = 0.89) models (**Table 2**, **Supplementary Figure S8A-C**). The molecules that contributed the most to the prediction of TB and LTBI were PC(14:0_22:6), PC(O-44:7), PC(O-24:1_22:6), PC(O-46:8) and PC(31:0), as well as clinical covariates of BMI (**Supplementary Figure S9A-C**). Remarkedly, the TB-ODx biosignature showed excellent performance in TB-LTBI prediction with AUCs of 0.94 in all applied algorithms (**Table 2**, **Supplementary Figure S8D-F**). PC(O-16:0_16:0), PC(O-18:1_16:0), PC(O-40:6), SM(34:1;2O), and FA(18:0) contributed significantly to the classification between TB and LTBI (**Supplementary Figure S9D-F**).

The capacity of the TB-NTM biosignature to predict TB-ODx was weaker than its performance in distinguishing TB from LTBI prediction, yet all AUCs > 0.68 in all three ML models (**Table 2**, **Supplementary Figure S10A-C**). RF models had the most variation in the AUCs compared to other models. PC(14:0_22:6), SM(39:2;2O), SM(35:1;2O), PC(O-36:1), and hypoxanthine were the most important metabolites for differentiating TB from ODx (**Supplementary Figure S11A-C**). The TB-LTBI biosignature exhibited good discriminatory power for classifying TB from ODx using RF, SVM, and PLS-DA models with AUC of 0.80, 0.79, and 0.83, respectively (**Table 2, Supplementary Figure S10D-F**). The most important features were SM(43:3;2O), FA(21:1), PC(16:0_16:0), PC(O-34:0), and PC(O-18:1_16:0) (Supplementary Figure S11D-F).

In the evaluation using the validation cohort, the TB-NTM biosignature showed an AUC of 0.72, 0.72, and 0.71 in three ML models of RF, SVM, and PLS-DA in differentiating TB from LTBI (**Table 3, Supplementary Figure S12A-C**). Similar results regarding the TB-LTBI and TB-ODx biosignatures were also demonstrated. Furthermore, the consensus biosignature exhibited an acceptable performance for classifying TB and LTBI, with the AUCs for RF, SVM, and PLS-DA models being 0.81, 0.71, and 0.72, respectively (**Supplementary Figure S12D)**. We also observed that in the discovery and validation cohorts, AUC obtained from the RF had higher variability and was less robust than the SVM and PLS-DA models.

**Table 3.** Performance of multi-ome biosignatures and clinical covariates for the differentiation of TB and LTBI in the validation cohort.

| Model | AUC ± SD | | | |
| --- | --- | --- | --- | --- |
|  | TB-NTM | TB-LTBI | TB-ODx | Consensus |
|  | Biosignature | Biosignature | Biosignature | Biosignature |
| **RF** | 0.72 ± 0.05 | 0.77 ± 0.08 | 0.71 ± 0.07 | 0.81 ± 0.09 |
| **SVM** | 0.72 ± 0.05 | 0.71 ± 0.03 | 0.68 ± 0.04 | 0.70 ± 0.02 |
| **PLS-DA** | 0.71 ± 0.04 | 0.73 ± 0.04 | 0.71 ± 0.01 | 0.76 ± 0.03 |

Abbreviation: AUC: Area under the receiver operating characteristic curve; LTBI: Latent tuberculosis infection; NTM: Nontuberculous mycobacteria; PLS-DA: Partial least squares discriminant analysis; ODx: Other diseases; TB: Tuberculosis; RF: Random forest; SD: Standard deviation; SVM: Support vector machines.

ML modeling using only the multi-ome biosignatures was conducted to evaluate the impact of clinical covariates and assess the potential performance of multi-ome biosignatures in discriminating TB from other non-TB groups. In both scenarios (i.e., with or without clinical covariates), the AUC (± SD) values were comparable across the tested cases in both the discovery and validation cohorts (**Tables 1, 2; Supplementary Tables S2, S3; Supplementary Figure S13-16**). Additionally, the RF model produced higher uncertainty and lower robustness in AUC compared with the SVM and PLS-DA models. The prominent omics-derived biomarkers remained consistent with those observed when clinical covariates were included (**Supplementary Figures S17)**.

### PC(14:0_22:6) is a promising biomarker for TB diagnosis

Among the ML-based classification analyses, PC(14:0_22:6) was consistently ranked as the most important predictor for differentiating active TB from other groups (**Supplementary Tables S4**). The abundance of PC(14:0_22:6) was consistently lower in the active TB group than in all other groups (**Figure 4A-C**). Additionally, PC(14:0_22:6) demonstrated a good performance (AUC = 0.73) in distinguishing between TB and LTBI in the validation cohort (**Figure 4C**).

**Figure 4.**
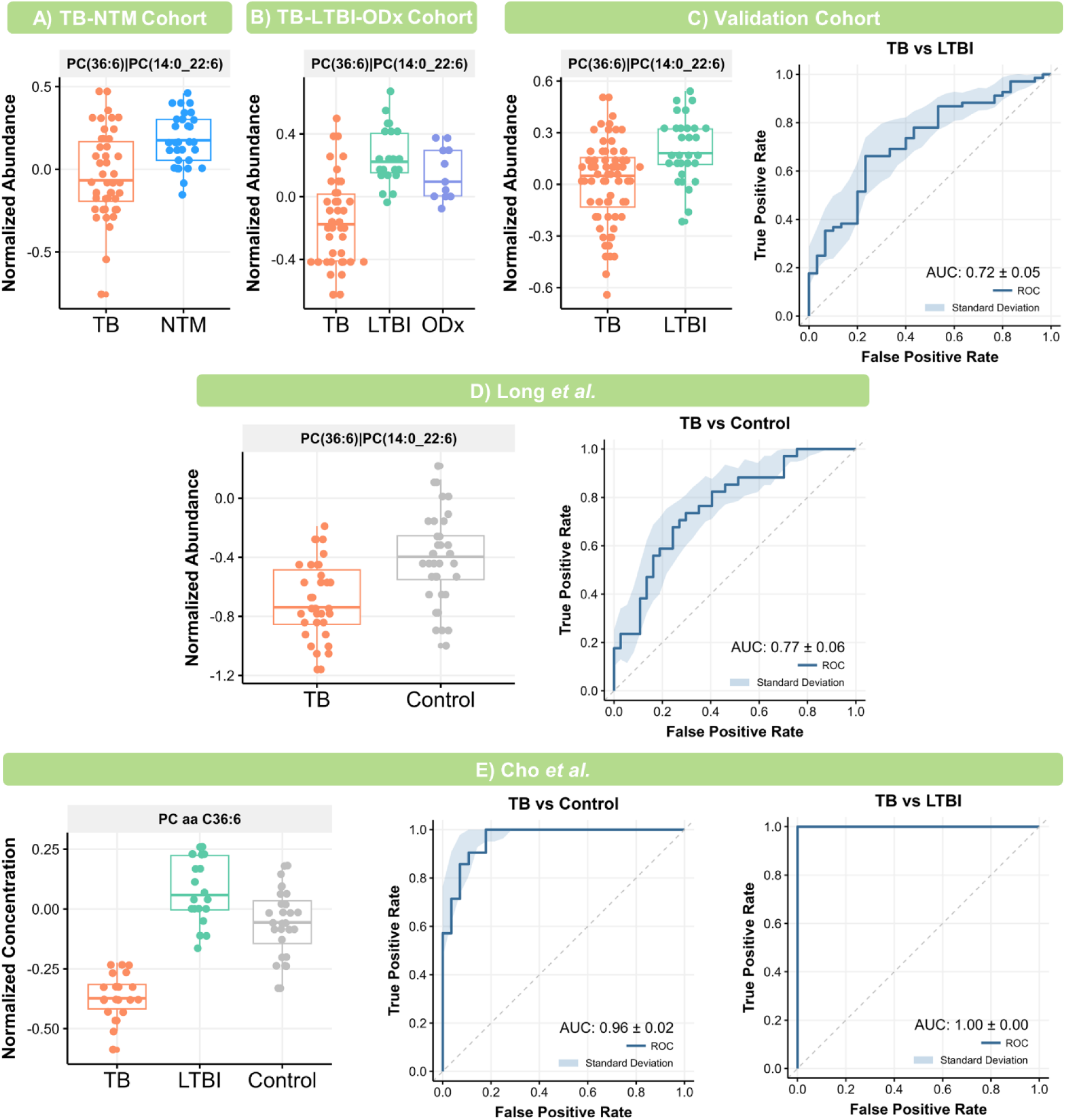
External validation of PC(14:0_22:6) as a TB diagnostic biomarker. **(A).** Relative abundance of PC(14:0_22:6) in the TB-NTM cohort. **(B)** Relative abundance of PC(14:0_22:6) in the TB-LTBI-ODx cohort. **(C)** Relative abundance of PC(14:0_22:6) and receiver operating characteristic (ROC) curve in the validation cohort. **(D)** Relative abundance of PC(14:0_22:6) and the ROC curve in the study of Long *et al*. **(E)** Normalized concentration of PC aa C36:6 and the ROC curves in the study of Cho *et al*.

Next, we conducted external validation to further evaluate the performance of PC(14:0_22:6) for TB diagnosis using data from our previously published lipidomics study [21]. The abundance of PC(14:0_22:6) was lower in TB patients than in the control group (**Figure 4D**). Univariate ROC curve analysis yielded an AUC of 0.76 for discriminating TB from controls. Additionally, we assessed the performance of PC(14:0_22:6) using quantitative data from a study by Cho *et al.* [31], under the assumption that PC(14:0_22:6) was a primary component of the reported lipid species PC aa C36:6. The TB group exhibited a significantly lower concentration of PC aa C36:6 compared with the LTBI and healthy control groups (**Figure 4E**). Notably, this lipid demonstrated excellent potential for distinguishing TB from LTBI (AUC = 1.00) and TB from health control (AUC = 0.97). Furthermore, univariate ROC analysis using unnormalized data resulted in AUC values ranging from 0.60 to 0.86 [21].

PC(14:0_22:6) also presented lower concentrations in the active TB group compared with non-TB controls (NTM, LTBI, and ODx) (**Figure 5A-C**). In univariate ROC analysis using concentration data across our three cohorts, the cross-validated AUC of PC(14:0_22:6) was 0.76 (± 0.03) for distinguishing active TB from non-TB controls (**Figure 5D**). Using a simple logistic regression model, PC(14:0_22:6) demonstrated similar effectiveness (i.e., AUC = 0.75 ± 0.04) in differentiating TB from non-TB groups (**Figure 5E**). However, the AUC significantly increased to 0.80, with less variability (SD = 0.02), when clinical covariates were included (**Figure 5F**). Overall, PC(14:0_22:6) shows promise as a biomarker for the differential diagnosis of TB from LTBI, NTM infections, and other lung diseases.

**Figure 5.**
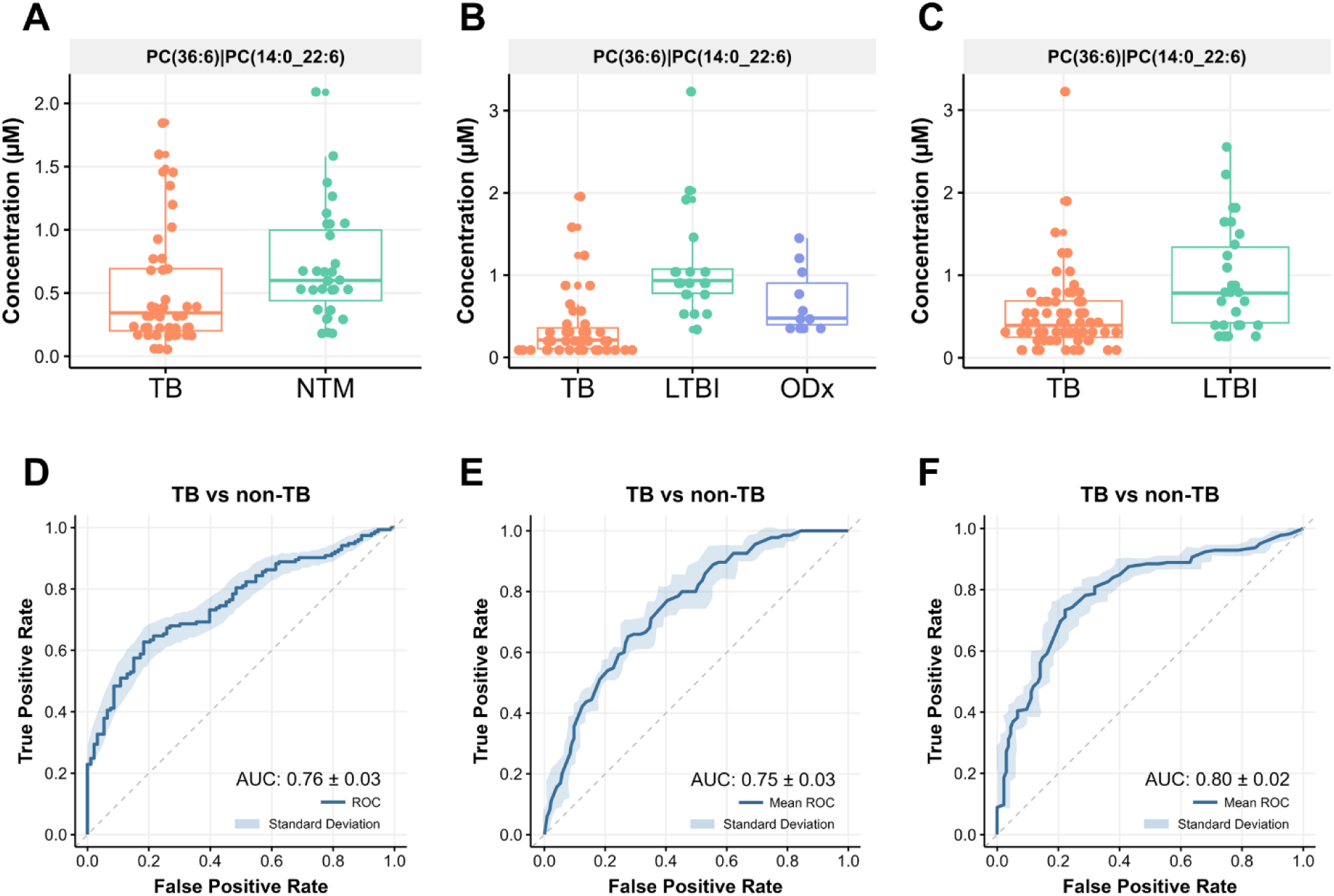
PC(14:0_22:6) concentration and receiver operating characteristic (ROC) curve analysis for the classification of TB and non-TB controls across the three cohorts. **(A)** Concentration in the TB-NTM cohort. **(B)** Concentration in the TB-LTBI-ODx cohort. **(C)** Concentration in the validation cohort. **(D)** Univariate ROC curve of the PC(14:0_22:6) concentration. **(E)** ROC curve obtained from logistic regression using PC(14:0_22:6) concentration without clinical covariates. **(F)** ROC curve obtained from logistic regression using PC(14:0_22:6) concentration with clinical covariates. Abbreviations: TB: tuberculosis; NTM: nontuberculous mycobacteria infection; LTBI: latent tuberculosis infection; and ODx: other lung diseases.

## DISCUSSION

This study utilized multi-omics coupled with ML methods to discover and validate plasma diagnostic biomarkers for TB across multiple cohorts. Two discovery cohorts and an independent validation cohort were used to derive several multi-ome biosignatures, which were then evaluated using predictive modeling for biomarker prioritization and external validation. Among different classes of biomarkers, lipids emerged as the most promising. The multi-ome biosignatures demonstrated good-to-excellent performance across groups in the discovery cohorts and showed adequate performance in the validation cohort. Notably, PC(14:0_22:6), which consistently presented lower levels in TB patients compared with non-TB controls, exhibited good predictability in external validation, establishing itself as a robust biomarker for TB diagnosis.

The development and use of biomarkers for TB diagnosis are crucial strategies to meet the WHO End TB targets by 2030 [14]. Although pathogen-derived biomarkers are commercially available and have been successfully implemented for TB diagnosis [14, 32], their detection efficacy is limited in paucibacillary specimens and often requires complex sampling from the disease site (e.g., sputum) [14, 33]. To address these limitations, the WHO has endorsed a paradigm shift towards the use of more easily obtainable samples, such as the host’s blood and urine [14]. To successfully translate biomarker discoveries into point-of-care tests, several strategies have been recommended. These include the design of studies with sufficient sample sizes and external validation, incorporation of established biological knowledge, and implementation of rigorous statistical and ML methodologies for biomarker discovery and validation [14, 33-36].

In a recent multi-national validation study evaluating 20 host transcriptome biosignatures for differentiating TB from ODx, none of the tested biosignatures met the WHO target product profiles (TPP) criteria when analyzing pooled data from six countries [37]. This underscores the ongoing need to explore clinically relevant biomarkers for TB diagnosis. State-of-the-art ML algorithms are valuable for biomarker discovery, evaluation, and prioritization [38]; however, the interpretability and biological plausibility of the predictive models are crucial [33]. In our study, predictive modeling with RF, SVM, and PLS-DA using three multi-omics-derived immunometabolic biosignatures demonstrated good-to-excellent performance across various scenarios. The multi-ome biosignatures also showed robust performance even without consideration of clinical covariates. Thus, our biosignatures provide biologically plausible and clinically relevant candidates for identifying the most promising biomarkers, facilitating external validation, and the translation of biomarkers into practical diagnostic tools [33].

PC(14:0_22:6) was proposed as a potential biomarker for TB diagnosis in our study. Although this lipid did not consistently meet the WHO TPP requirements in some of our validations (with AUCs ranging from 0.80 to 1.00), a diagnostic test that includes clinical covariates and combines PC(14:0_22:6) with other biomarkers could enhance the accuracy of diagnosing actual TB cases [33]. Notably, consistent with our findings, studies by Han *et al.* and Chen *et al.* also observed a decrease in PC(14:0_22:6) levels in TB patients and highlighted its potential to differentiate TB from healthy individuals and other control groups [39, 40]. The performance of the proposed biomarkers in our study has been lower because it was also tested against NTM, a condition with significant overlapping manifestations with active TB.

Host-circulating lipids have been reported as potential biomarkers for diagnosing TB [31, 40, 41]. Notably, our previous work consistently highlighted the role of lipids in the pathogenesis of TB and their potential to aid in TB management [21, 42]. We also found that host metabolic perturbations in TB were associated with innate immune responses to the pathogen [22]. PC has been implicated in the host immune response to pathogens [21, 42, 43], and PC-derived lipid mediators play a role in immune modulation [44]. This supports the rationale for using PC-sourced molecules as diagnostic biomarkers for TB. Additionally, *Mtb* has been reported to utilize host TGs to form lipid droplets inside macrophages, serving as a nutrient source [40]. We identified declining levels of TGs in TB patients relative to those with LTBI and ODx, which may be attributed to the *Mtb* consumption. Intriguingly, TG levels in NTM infections were even lower than those observed in TB, as revealed by both conventional and integrative analyses. This finding could suggest that NTM also utilizes host TGs to form lipid droplets [45] or that the lower levels of TGs in NTM might result from complex metabolic interactions between the pathogen and the host.

Our study, using plasma samples, revealed minimal differences in the hydrophilic metabolome and immune profile between active TB and non-TB controls. Notably, tryptophan metabolism-related metabolites emerged as significant for TB differential diagnosis. Our previous research also identified alterations in tryptophan metabolism between TB and NTM by analyzing the urinary metabolome [46]. Additionally, citrate, a metabolite associated with the TCA cycle, showed potential in aiding the diagnosis of TB versus NTM and TB versus LTBI in our study. Although there is limited evidence supporting citrate as a diagnostic biomarker for TB, the TCA cycle plays a crucial role in bacterial growth and inflammatory responses in pulmonary TB patients [47]. Among immune factors, VEGF-A demonstrated significance in distinguishing TB from NTM but not TB from LTBI, which contrasted with previous findings [19]. Other notable analytes (hypoxanthine and IL-15) were consistently found at lower levels in TB patients than in non-TB controls across all three biosignatures. However, their contributions to differentiating TB from non-TB groups were minimal.

Although our multi-cohort study design allowed for comprehensive analysis, the small and imbalanced sample sizes in each cohort could limit the power to identify biomarkers with subtle but robust differences between TB and non-TB controls. Nonetheless, we validated the cohort-specific biosignatures across our cohorts and other external populations. Further studies are needed to confirm the proposed biomarker in different clinical settings. Additionally, a technical limitation of our study is the lack of absolute quantification for complex lipids, largely due to the absence of suitable authentic standards. Further research should focus on developing reliable assays that are more appropriate for clinical implementation. Finally, an integrative analysis incorporating blood transcriptomics and plasma proteomics would provide a more comprehensive immunometabolic landscape of TB, NTM, LTBI, and ODx. There is a significant yet unmet need for further mechanistic elucidation using a comprehensive multi-omics approach.

## CONCLUSION

We identified perturbations in the immune, metabolome, and lipidome profiles associated with TB rather than NTM, LTBI, and ODx. We derived four multi-ome biosignatures, which demonstrated strong performance in classifying active TB versus non-TB controls. Predictive modeling highlights lipids as potential biomarkers for differentiating TB from NTM and for distinguishing TB from LTBI and ODx. Further prospective multi-center studies are needed to validate PC(14:0_22:6) as a diagnostic biomarker for tuberculosis management. In conclusion, our study demonstrated that the integration of multi-omics analysis with predictive ML modeling is effective for discovering and validating robust biomarkers for distinguishing TB from other complex conditions with overlapping clinical manifestations.

## Data Availability

The data supporting this study's findings are available upon reasonable request.

## CRediT authorship contribution statement

**Nguyen Tran Nam Tien**: Data curation, Methodology, Investigation, Formal analysis, Visualization, Validation, Writing – Original Draft, Writing – Review & Editing. **Nguyen Thi Hai Yen**: Data curation, Methodology, Investigation, Formal analysis, Visualization, Validation, Writing – Original Draft, Writing – Review & Editing. **Nguyen Ky Phat**: Data curation, Methodology, Formal analysis, Visualization, Validation, Writing – Review & Editing. **Nguyen Ky Anh:** Data curation, Methodology, Software, Validation, Writing – Original Draft, Writing – Review & Editing. **Nguyen Quang Thu**: Data curation, Methodology, Software, Validation, Writing – Review & Editing. **Cho Eunsu**: Data curation, Methodology, Software, Writing – Review & Editing. **Ho-Sook Kim**: Methodology, Validation, Resources, Writing – Review & Editing. **Vu Dinh Hoa:** Methodology, Validation, Writing – Review & Editing. **Duc Ninh Nguyen:** Methodology, Validation, Writing – Review & Editing. **Dong Hyun Kim**: Conceptualization, Validation, Supervision, Resources, Writing – Review & Editing, Funding acquisition. **Jee Youn Oh**: Conceptualization, Methodology, Investigation, Validation, Resources, Supervision, Writing – Review & Editing, Funding acquisition. **Nguyen Phuoc Long**: Conceptualization, Methodology, Investigation, Formal analysis, Validation, Resources, Supervision, Writing – Original draft, Writing – Review & Editing, Funding acquisition.

## Acknowledgments

We thank Tam Anh Trinh and Da Dat Ly for carrying out the immune profiling assay.

