## Supplementary Materials for "Circulating Lipids as Biomarkers for Diagnosis of Tuberculosis: A Multi-cohort, Multi-omics Data Integration Analysis"

### SUPPLEMENTARY METHODS

#### Chemicals, reagents, and consumables

The LC-MS grade solvents [water, methanol (MeOH), acetonitrile (ACN), and isopropanol (IPA)] were obtained from Merck KGaA (Darmstadt, Germany). Reagents [formic acid, ammonium acetate, ammonium formate, methyl *tert*-butyl ether (MTBE), and toluene] as well as Mass Spectrometry Metabolite Library of Standards (msmls-lot-230-01) were provided by Sigma-Aldrich (St. Louis, Missouri, USA). For metabolomics experiment, the internal standards (IS) including acetyl-L-carnitine-(N-methyl-d3), L-phenyl-d5-alanine, L-tryptophan-(indole-d5), leucine enkephalin, SM(d18:1/15:0)-d9 and cholic acid-2,2,3,4,4-d5 were acquired from Sigma-Aldrich (St. Louis, Missouri, USA). The ACQUITY UPLC HSS T3 column (50 mm × 2.1 mm; 1.8 µm) coupled with ACQUITY UPLC HSS T3 VanGuard pre-column (5 mm × 2.1 mm; 1.8 µm) was utilized (Waters, Milford, MA, USA). For untargeted lipidomics, the IS of SPLASH® LIPIDOMIX® Mass Spec Standard and Deuterated Ceramide LIPIDOMIX® Mass Spec Standard was purchased from Avanti Polar Lipids (Alabama, USA). The ACQUITY UPLC BEH C18 column (50 mm × 2.1 mm; 1.7 µm) linked to ACQUITY UPLC BEH C18 VanGuard pre-column (5 mm × 2.1 mm; 1.7 µm) was used (Waters, Milford, MA, USA). In the immune profiling experiment, the customized 40-analyte Multiplex kits (MILLIPLEX® Human Cytokine/Chemokine/Growth Factor Panel A − Immunology Multiplex Assay, HCYTA-60K) were purchased from Merck-Millipore (MA, United States). Reagents (i.e., cytokine/chemokine/growth Factor standards and quality controls, assay buffer, wash buffer, detection antibodies, streptavidin-phycoerythrin, and bead diluent) for immunoassay were provided simultaneously with the kits.

#### Metabolomics sample treatment and instrumental analysis

The protocol from previous works with modifications was applied to extract metabolite from the plasma sample [1-5]. Briefly, we used ice to thaw a 20 μL sample (−80 °C) for about 30 min. All samples were then vortexed for 10 s. Next, 80 μL of 6 ISs pre-dissolved MeOH (−80 °C) was added to each sample. In the next step, 30 s vortex prior to 2 min centrifugation (14,000 *rcf* and 4 ºC) was applied to obtain 70 μL supernatant, which then was evaporated with a nitrogen gas system. The dried extracts were stored at -80 ºC until analysis. Consequently, 200 μL of 50% MeOH (4 ºC) was used for resuspending the dried samples. The samples were then vortexed vigorously and centrifuged at 14,000 *rcf* and 4 ºC for 2 min to obtain supernatant. A volume of 150 μL was used for LC-MS data acquisition, and 20 μL of each sample was gathered to make a pooled quality control (QC) sample. The samples were stored at an autosampler at 4 ºC.

The apparatus of the Shimadzu Nexera LC system (Kyoto, Japan) and the X500R Quadrupole Time-of-Flight mass spectrometer combined with Turbo V™ ion source and TwinSpray probe (SCIEX, MA, USA) was implemented. The stationary phase was the ACQUITY UPLC HSS T3 column (50 mm × 2.1 mm; 1.8 μm) coupled with ACQUITY UPLC HSS T3 VanGuard pre-column (5 mm × 2.1 mm; 1.8 μm). Mobile phase A was 100% water with 0.2% formic acid and mobile phase B was 100% ACN with 0.1% formic acid. The information-dependent mode was used to acquire the data with a 2 μL injection volume for both positive ion mode (POS) and negative ion mode (NEG). For POS, 2 μL was eventually used after exploring the results of 1 μL injection. After every eight injections, mass calibration by the X500R built-in calibrant delivery system was conducted.

**Lipidomics sample treatment and instrumental analysis**

The lipid extraction from plasma followed the established procedures with some adjustments [1-4, 6]. First, 20 μL plasma samples (−80 °C) were thawed on ice for about 30 min.

A prepared IS mixture of SPLASH and Deuterated Ceramide LIPIDOMIX (4 μL, 1:1, *v/v*) was pipetted to each sample, followed by 10-s vortexing. The mixture was then incubated on ice for 20 min and vortexed briefly. Afterward, 300 μL of MeOH (−20 °C) and 1000 μL of MTBE (−20 °C) were added and vortexed briefly. The mixtures were incubated in a shaker (20 min at 1200 *rpm* and 4 °C). Afterward, 250 μL of water (4 °C) was put in, vortexed for 20 s, followed by 10-min incubation on ice. Next, the samples were centrifuged (2 min, 4 °C, 14,000 *rcf*) to collect 500 μL of upper layer to dry using a nitrogen gas flow. The dried extracts were stored at -80 °C until analysis. We used 200 μL MeOH/toluene (4 °C, 9:1, *v/v*) to resuspend the dried samples, followed by 2-min centrifugation (4 °C and 14,000 *rcf*) to acquire 150 μL of supernatant for the LC-MS injection. Besides, 20 μL of supernatant from each sample was used to create the post-extraction pooled QC sample. The samples were stored at an autosampler at 4 °C.

The LC-MS instrument was similar to the untargeted metabolomics experiments. The ACQUITY UPLC BEH C18 column (50 mm × 2.1 mm; 1.7 μm) connected to an ACQUITY UPLC BEH C18 VanGuard pre-column (5 mm × 2.1 mm; 1.7 μm) was implemented. In the POS, mobile phase A was ACN/water (60:40, *v/v*) with 10 mM ammonium formate and 0.1% formic acid and mobile phase B was IPA/ACN (90:10, *v/v*) containing 10 mM ammonium formate and 0.1% formic acid. For the NEG, mobile phase A was ACN/water (60:40, *v/v*) with 10 mM ammonium acetate and mobile phase B was IPA/ACN (90:10, *v/v*) containing 10 mM ammonium acetate. A volume of 2 μL for POS and 3 μL for NEG injection were applied. During the analysis procedure, eight injection cycles of mass calibration and data-dependent mode were carried out.

**Immune profiling sample treatment and instrumental analysis**

The concentrations of 40 cytokines were simultaneously quantified with the Luminex ® 200™ system (Luminex Corporation, Austin, TX, United States). The experiment was followed strictly by the manufacturer's protocol. Briefly, the multiplex kit and reagents were warmed to room temperature for at least 30 min before conducting the experiment. Samples (-80 ºC) were first thawed and vortexed before performing the experiment. In the 96-well plate, plasma sample wells were prepared by using 25 µL of plasma sample and 25 µL of assay buffer. For the standard, quality control, and background wells, 25 µL of each corresponding solution followed by 25 µL of the matrix solution were used. Next, 40 groups of analyte-conjugated beads were mixed and dispensed at a volume of 25 µL per well into each well. This mixture was then incubated at room temperature for 2 h under a shaking condition at 500 *rpm* using an orbital shaker (MixM-1500, Daihan Scientific, South Korea). After the incubation, the plate underwent three rounds of washing using the Biotek 405 TS washer (Merck-Millipore, MA, United States). Afterward, we added 25 µL of detection antibodies to each well. The mixture was then incubated at room temperature (1 h under shaking conditions at 500 *rpm* using an orbital shaker), followed by a three-time washing step. Next, 25 µL of streptavidin-phycoerythrin solution was put in and further incubated for 30 min at room temperature with shaking at 500 *rpm*. After another three rounds of washing, 150 µL sheath fluid buffer was added. The plate was shaken at 500 *rpm* for 5 min at room temperature. Finally, the plate was read using a Luminex 200$^{TM}$ instrument. In this study, no plasma samples had a cytokine concentration above the maximum detectable concentration, so the dilution was unnecessary.

**Data preprocessing and data treatment**

The data-dependent module in MS-DIAL (version 4.9.0) was used to process the MS data. Our authentic standards in-house libraries and MS-DIAL in-built and public libraries were employed for automated annotation and manual annotation inspection by at least two researchers [2, 7-9]. The NEG and POS ion modes were combined. Annotated features with 2 or more peaks or being detected in two ion modes were subjected to manual exploration. This process yielded robustly and uniquely annotated analytes for downstream analyses. Utilizing MetaboAnalyst 6.0, features with a missing rate ≥ 50% were excluded, whereas the k-nearest neighbors-based imputation method was used for the retained missing values [10]. The features with relative standard deviation ≥ 25% in the QCs samples were also excluded.

The Belysa® software (Merck-Millipore, MA, United States) was used to establish the calibration curve for immune profiling data. The automatic optimization fitting procedure was used with appropriate weighting parameters. Statistical outliers of the standards were also detected and removed during the curve fitting procedure. Eventually, the four- and five-parameter logistic regression with $1/y^2$ weighting was the majority. Interpolated and extrapolated concentrations over the limit of detection (LoD) were used to select the cytokines for analyses. Analytes whose ≥ 50% samples were recorded as not detectable (ND) and whose ≥ 50% extrapolated concentrations were excluded. This resulted in 8 and 7 analytes being removed in the TB-NTM and TB-LTBI-ODx cohorts, respectively. For subsequent analyses, the median fluorescence intensity (MFI) values were used [11].

### SUPPLEMENTARY FIGURES

**Figure S1. Principal components analysis scores plots in the discovery cohorts. (A)** Immune profiles between TB and NTM. **(B)** Metabolite profiles between TB and NTM. **(C)** Lipid profiles between TB and NTM. **(D)** Immune profiles between TB and LTBI. **(E)** Metabolite profiles between TB and LTBI. **(F)** Lipid profiles between TB and LTBI. **(G)** Immune profiles between TB and ODx. **(H)** Metabolite profiles between TB and ODx. **(I)** Lipid profiles between TB and ODx. Abbreviations: TB: tuberculosis; NTM: nontuberculous mycobacteria infection; LTBI: latent tuberculosis infection; and ODx: other lung diseases.

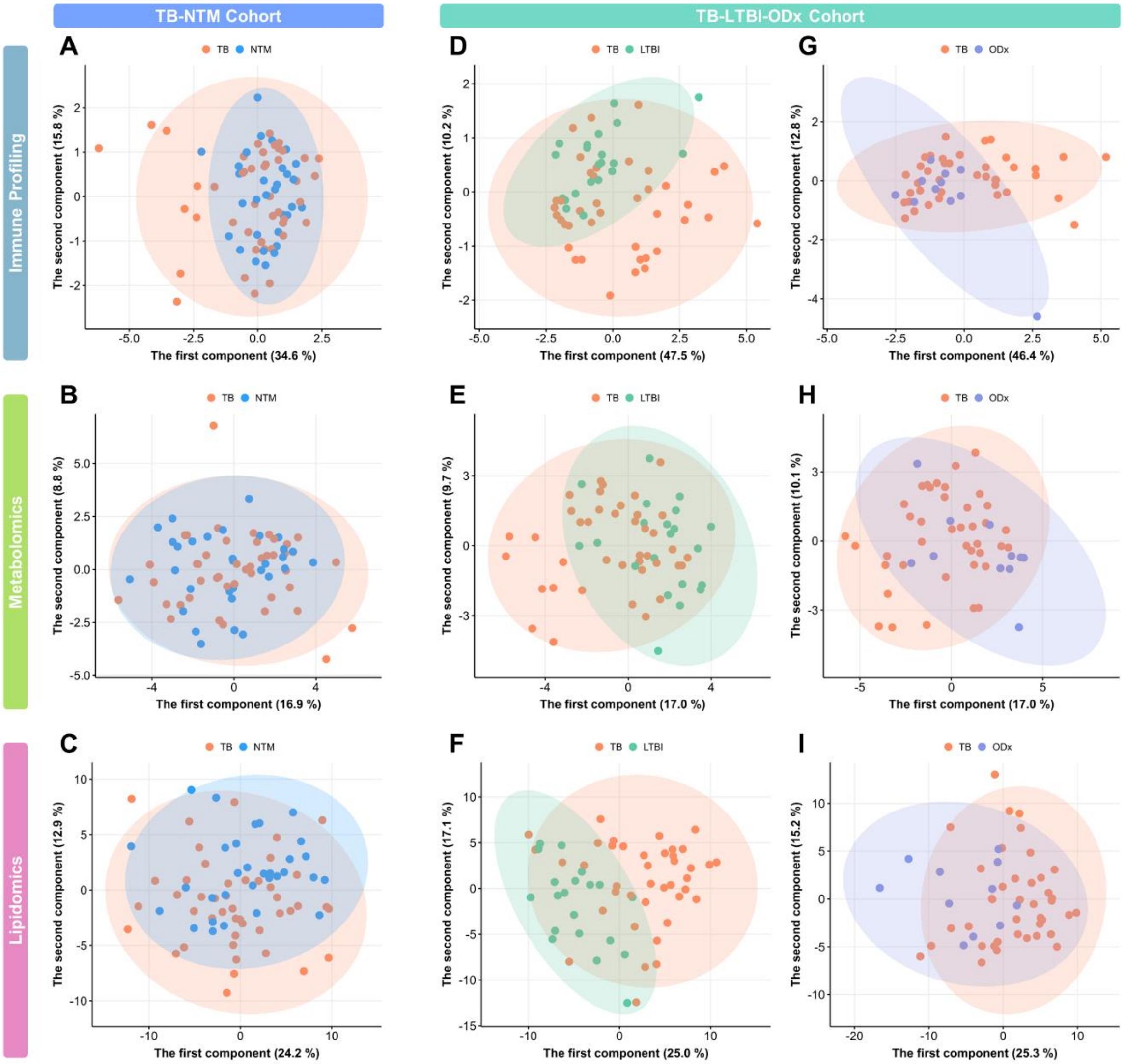

**Figure S2. DIABLO diagnostic plot and receiver operating characteristic (ROC) curve for each data layer using all components in the TB-NTM cohort. (A)** Diagnostic plot of DIABLO-selected biosignatures in component 1. Three left panels show pair-wise correlation coefficients. Three right panels present samples colored by groups with their 95% confidence ellipse. **(B**) ROC curve for the immune profiling data. **(C)** ROC curve for the metabolomics data. **(D**) ROC curve for the lipidomics data.

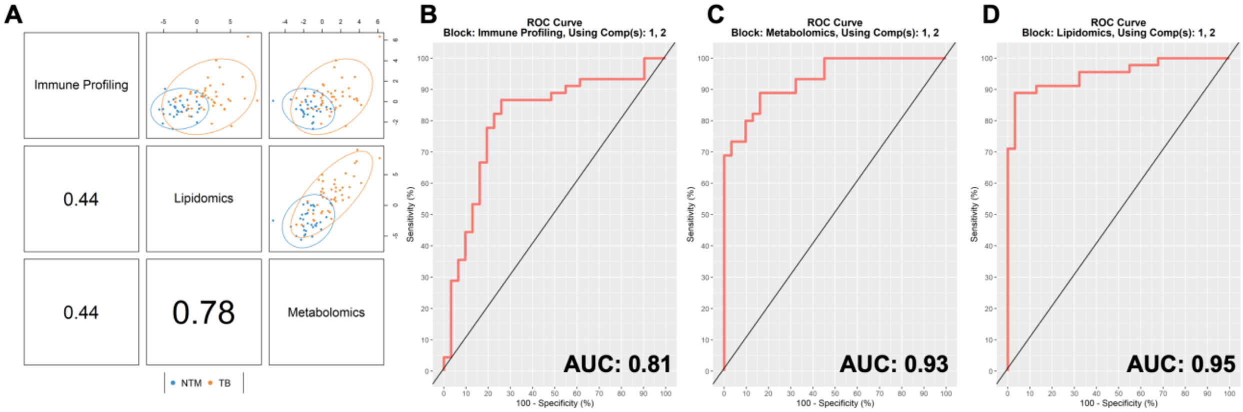

**Figure S3. DIABLO diagnostic plot and receiver operating characteristic (ROC) curve for each data layer using all components in the TB-LTBI-ODx cohort for TB versus LTBI comparison. (A)** Diagnostic plot of DIABLO-selected biosignatures in component 1. Three left panels show pair-wise correlation coefficients. Three right panels present samples colored by groups with their 95% confidence ellipse. **(B)** ROC curve for the immune profiling data. **(C)** ROC curve for the metabolomics data. **(D)** ROC curve for the lipidomics data.

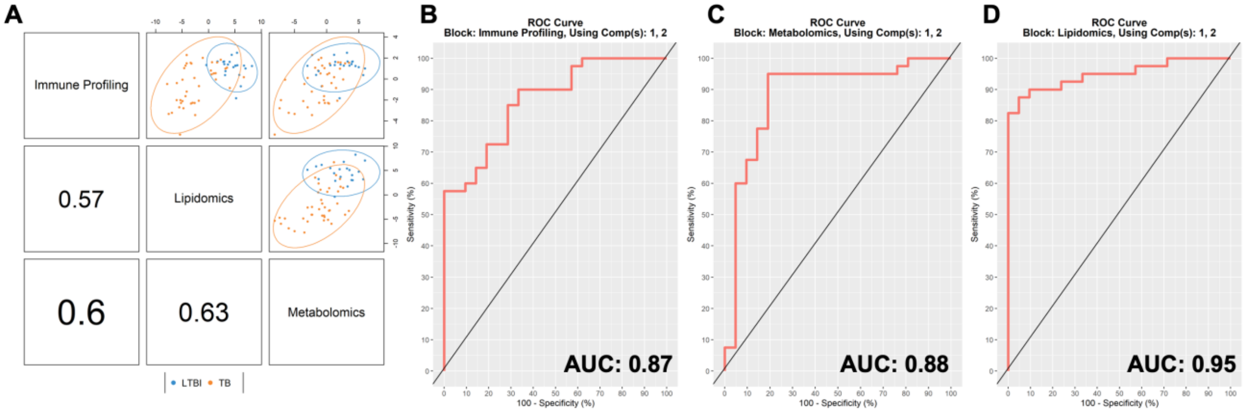

**Figure S4. DIABLO diagnostic plot and receiver operating characteristic (ROC) curve for each data layer using all components in the TB-LTBI-ODx cohort for TB versus ODx comparison. (A)** Diagnostic plot of DIABLO-selected biosignatures in component 1. Three left panels show pair-wise correlation coefficients. Three right panels present samples colored by groups with their 95% confidence ellipse. **(B)** ROC curve for the immune profiling data. **(C)** ROC curve for the metabolomics data. **(D)** ROC curve for the lipidomics data.

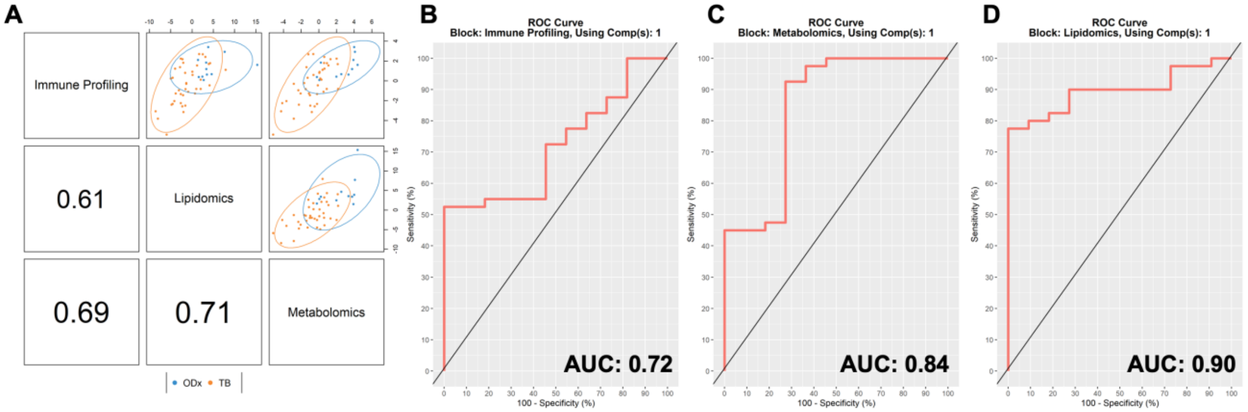

**Figure S5. The intersection of the three multi-ome biosignatures to establish the consensus biosignature consisting of shared biomarkers in at least 2/3 biosignatures.**

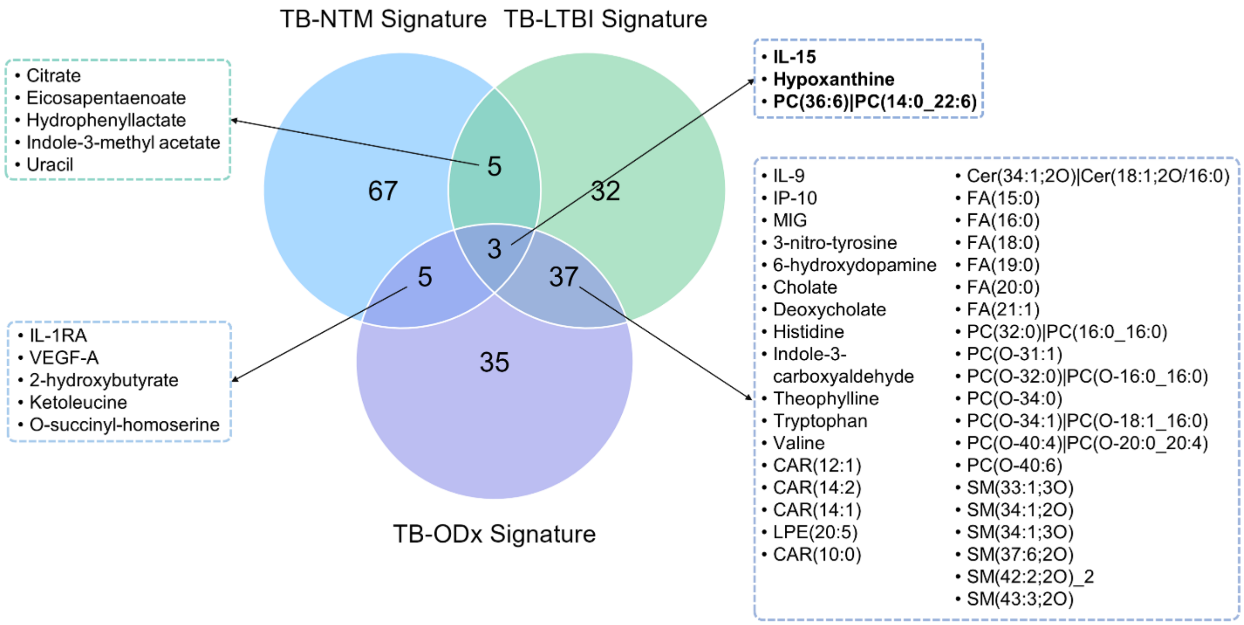

**Figure S6. Receiver operating characteristic curve of the random forest (RF), support vector machines (SVM), and partial least squares discriminant analysis (PLS-DA) models using the TB-LTBI biosignature (A-C) or the TB-ODx biosignature (D-F), with clinical covariates, for the classification of TB and NTM.** Abbreviations: TB: tuberculosis; NTM: nontuberculous mycobacteria infection; LTBI: latent tuberculosis infection; and ODx: other lung diseases.

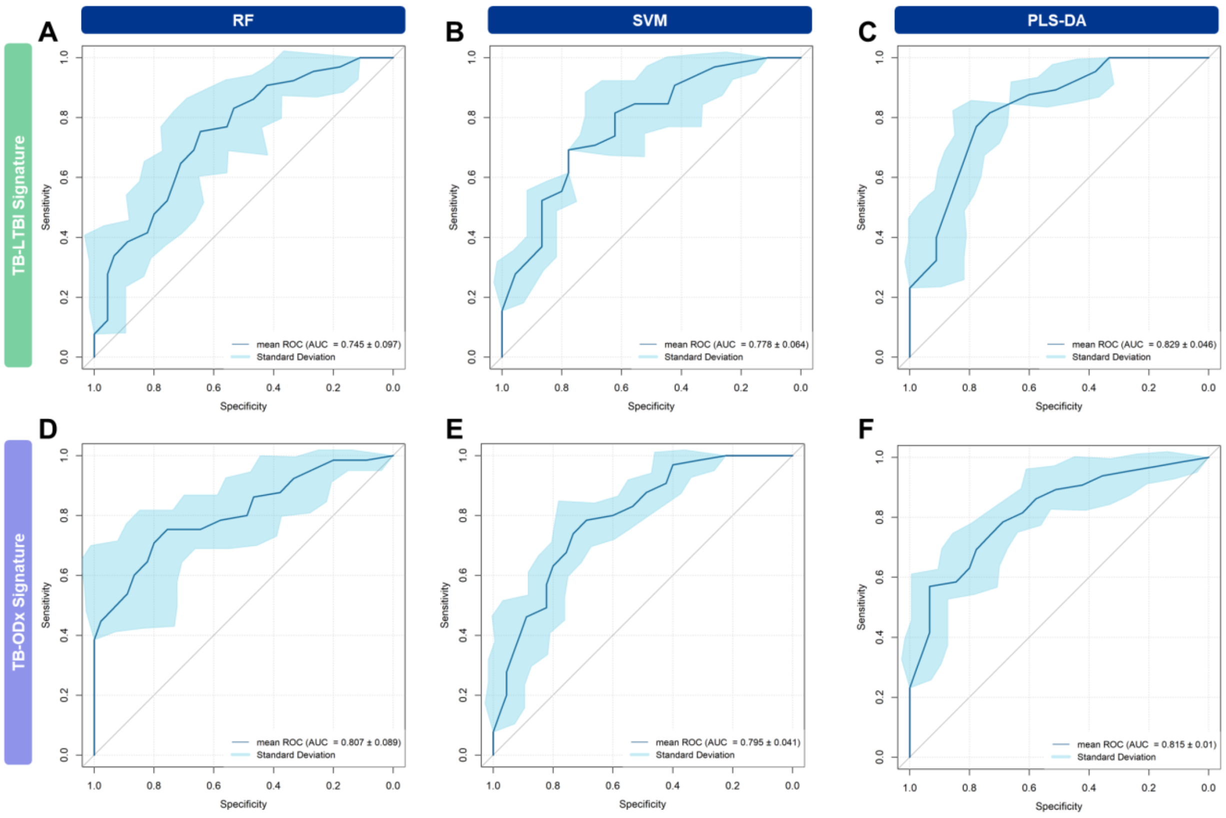

**Figure S7. Top 10% variables based on importance score of models for the classification of TB and NTM.** **(A)** RF with TB-LTBI biosignature and clinical covariates. **(B)** SVM with TB-LTBI biosignature and clinical covariates. **(C)** PLS-DA with TB-LTBI biosignature and clinical covariates. **(D)** RF with TB-ODx biosignature and clinical covariates. **(E)** SVM with TB-ODx biosignature and clinical covariates. **(F)** PLS-DA with TB-ODx biosignature and clinical covariates. Abbreviations: TB: tuberculosis; NTM: nontuberculous mycobacteria infection; LTBI: latent tuberculosis infection; and ODx: other lung diseases; RF: random forest; SVM: support vector machines; PLS-DA: partial least squares discriminant analysis.

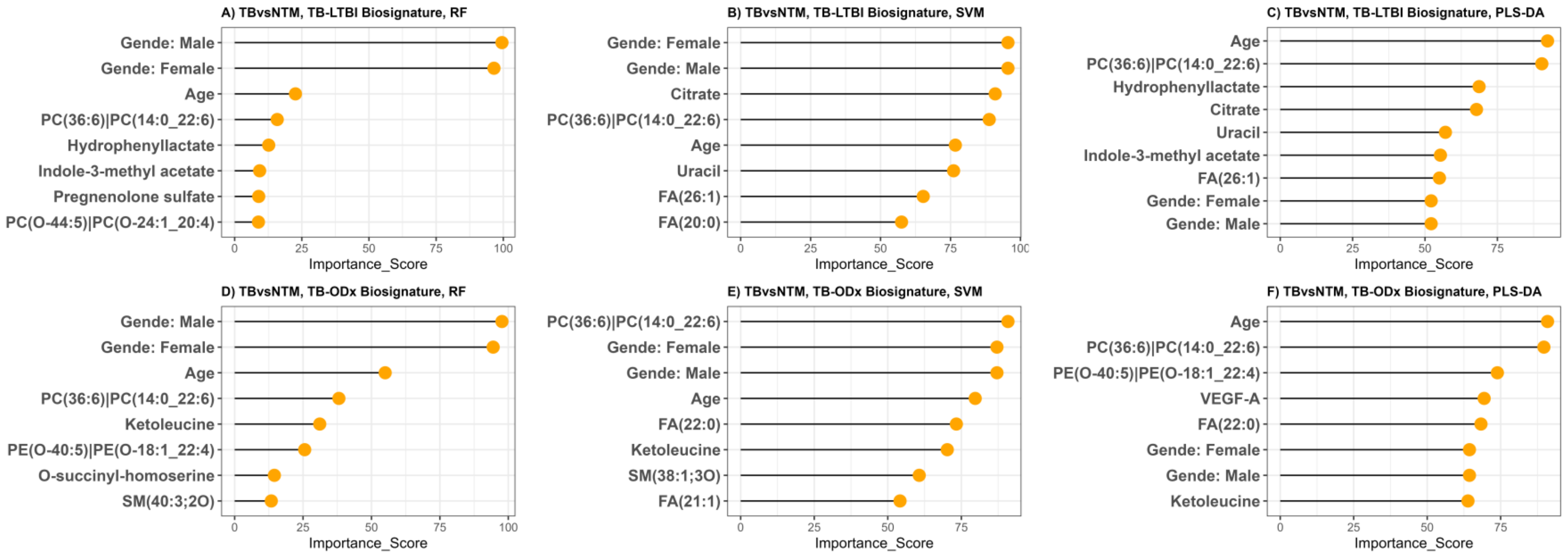

**Figure S8. Receiver operating characteristic curve of the random forest (RF), support vector machines (SVM), and partial least squares discriminant analysis (PLS-DA) models using the TB-NTM biosignature (A-C) or the TB-ODx biosignature (D-F), with clinical covariates, for the classification of TB and LTBI.** Abbreviations: TB: tuberculosis; NTM: nontuberculous mycobacteria infection; LTBI: latent tuberculosis infection; and ODx: other lung diseases.

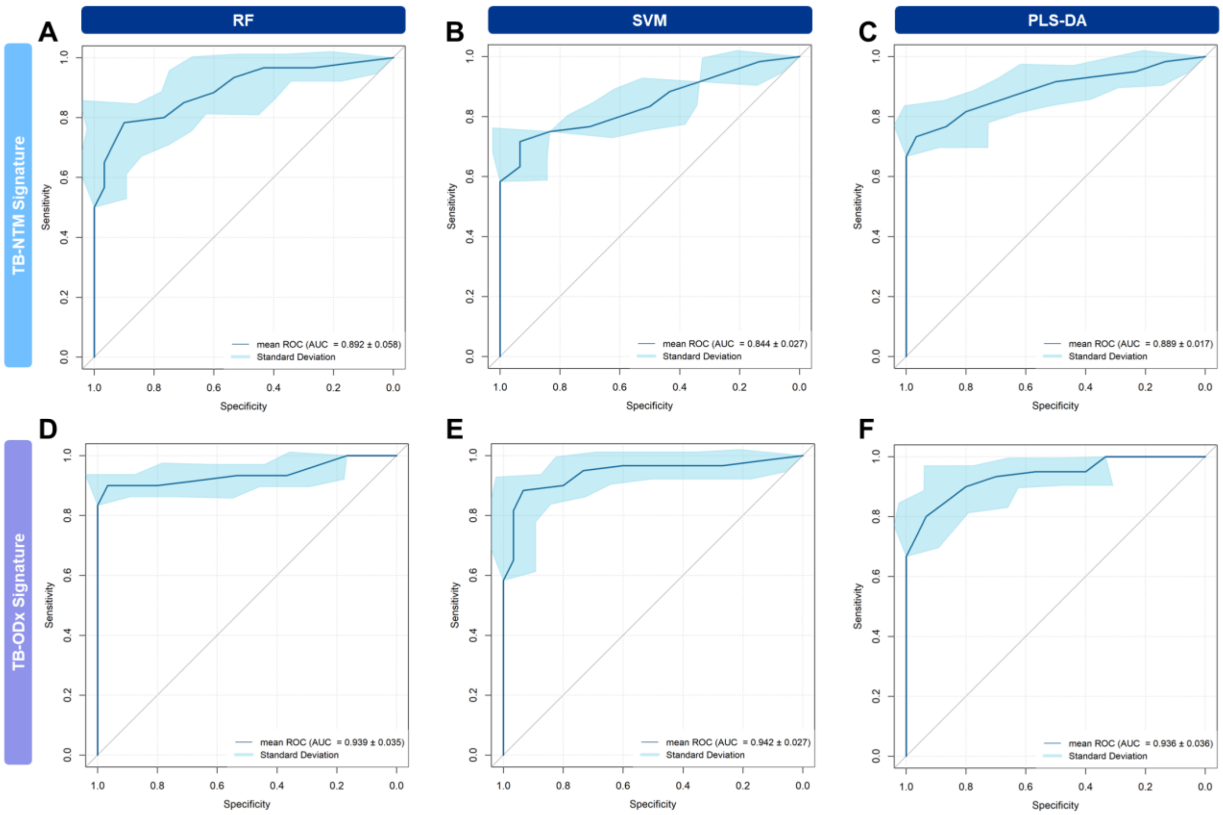

**Figure S9. Top 10% variables based on importance score of models for the classification of TB and LTBI. (A)** RF with TB-NTM biosignature and clinical covariates. **(B)** SVM with TB-NTM biosignature and clinical covariates. **(C)** PLS-DA with TB-NTM biosignature and clinical covariates. **(D)** RF with TB-LTBI biosignature and clinical covariates. **(E)** SVM with TB-LTBI biosignature and clinical covariates. **(F)** PLS-DA with TB-LTBI biosignature and clinical covariates. Abbreviations: TB: tuberculosis; NTM: nontuberculous mycobacteria infection; LTBI: latent tuberculosis infection; and ODx: other lung diseases; RF: random forest; SVM: support vector machines; PLS-DA: partial least squares discriminant analysis.

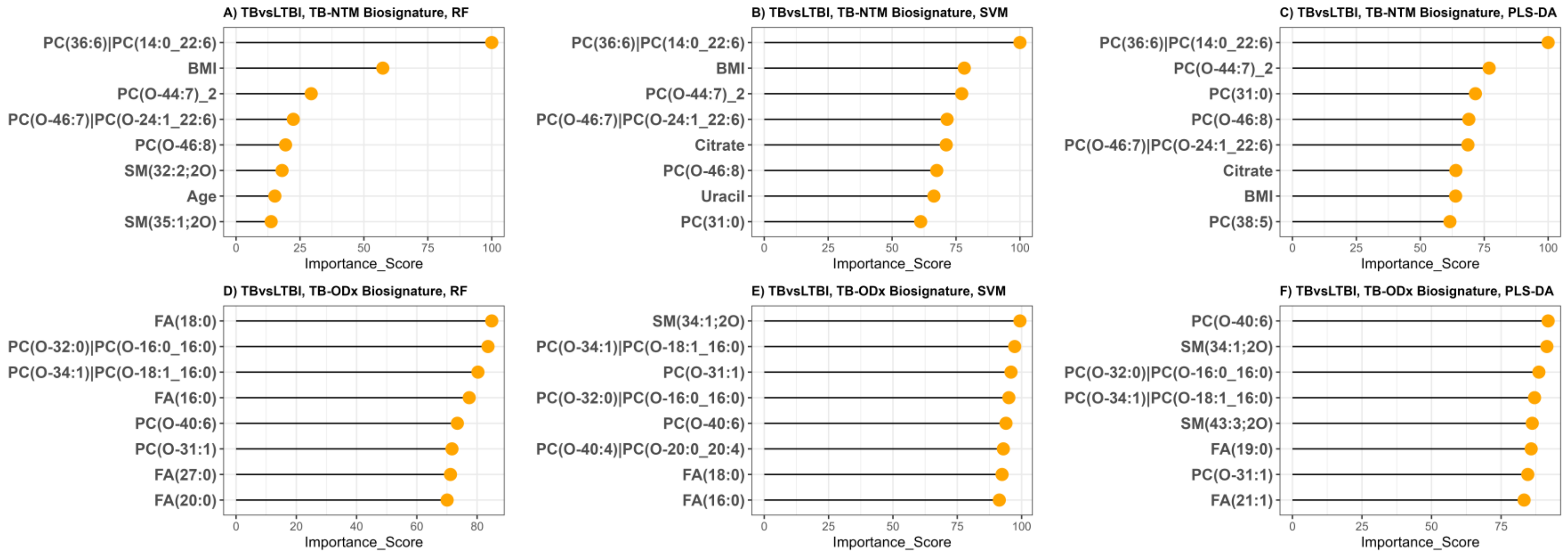

**Figure S10. Receiver operating characteristic curve of the random forest (RF), support vector machines (SVM), and partial least squares discriminant analysis (PLS-DA) models using the TB-LTBI biosignature (A-C) and the TB-ODx biosignature (D-F), with clinical covariates, for the classification of TB and ODx.** Abbreviations: TB: tuberculosis; NTM: nontuberculous mycobacteria infection; LTBI: latent tuberculosis infection; and ODx: other lung diseases.

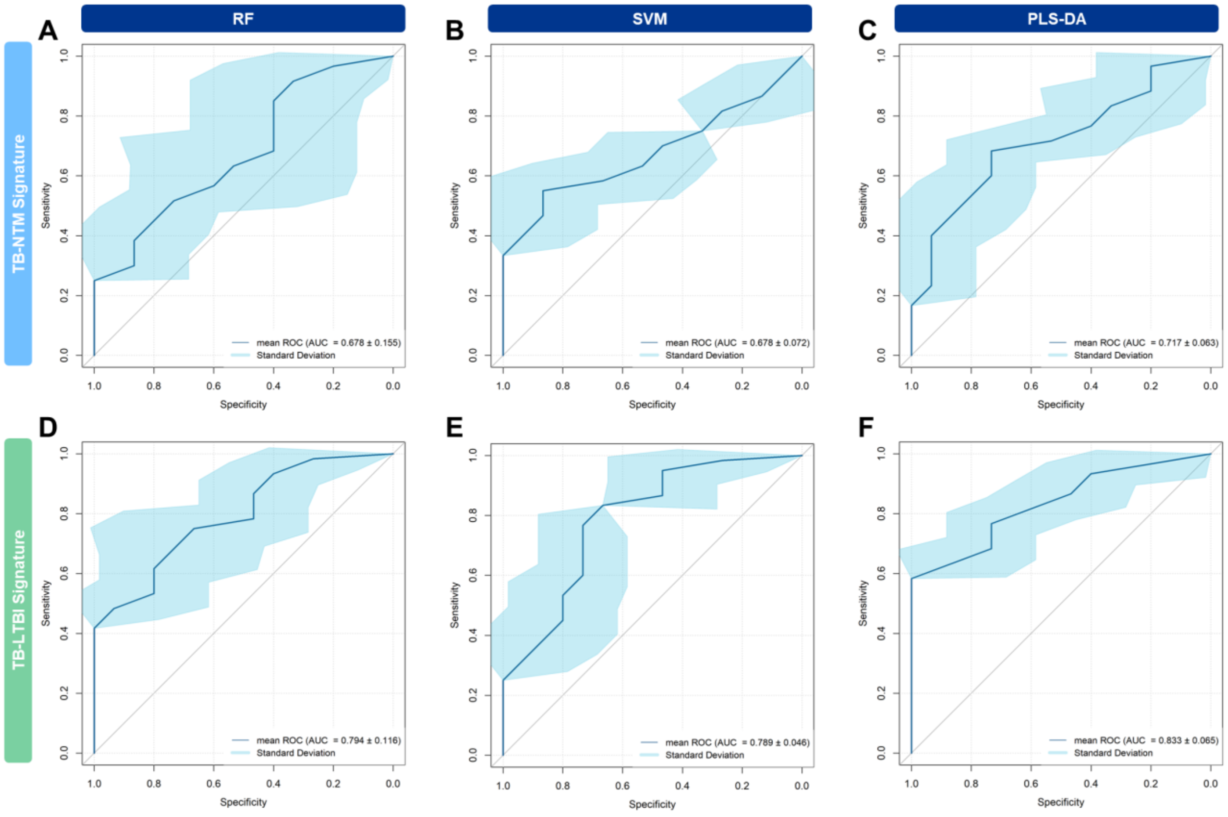

**Figure S11. Top 10% variables based on importance score of models for the classification of TB and ODx.** **(A)** RF with TB-NTM biosignature and clinical covariates. **(B)** SVM with TB-NTM biosignature and clinical covariates. **(C)** PLS-DA with TB-NTM biosignature and clinical covariates. **(D)** RF with TB-ODx biosignature and clinical covariates. **(E)** SVM with TB-ODx biosignature and clinical covariates. **(F)** PLS-DA with TB-ODx biosignature and clinical covariates. Abbreviations: TB: tuberculosis; NTM: nontuberculous mycobacteria infection; LTBI: latent tuberculosis infection; and ODx: other lung diseases; RF: random forest; SVM: support vector machines; PLS-DA: partial least squares discriminant analysis.

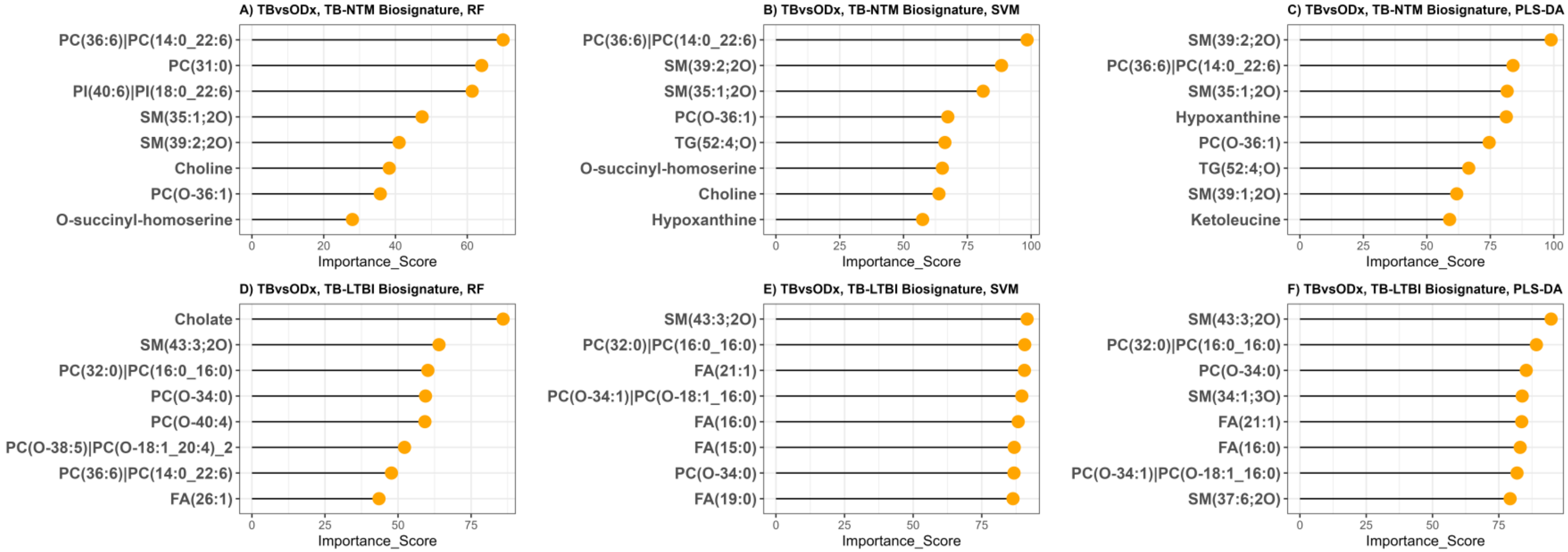

**Figure S12. Receiver operating characteristic curve of the random forest (RF), support vector machines (SVM), and partial least squares discriminant analysis (PLS-DA) models using the TB-NTM biosignature (A), the TB-LTBI biosignature (B), the TB-ODx biosignature (C), or the consensus biosignature (D), with clinical covariates, for the differentiation of TB and LTBI in the validation cohort.** Abbreviations: TB: tuberculosis; NTM: nontuberculous mycobacteria infection; LTBI: latent tuberculosis infection; and ODx: other lung diseases.

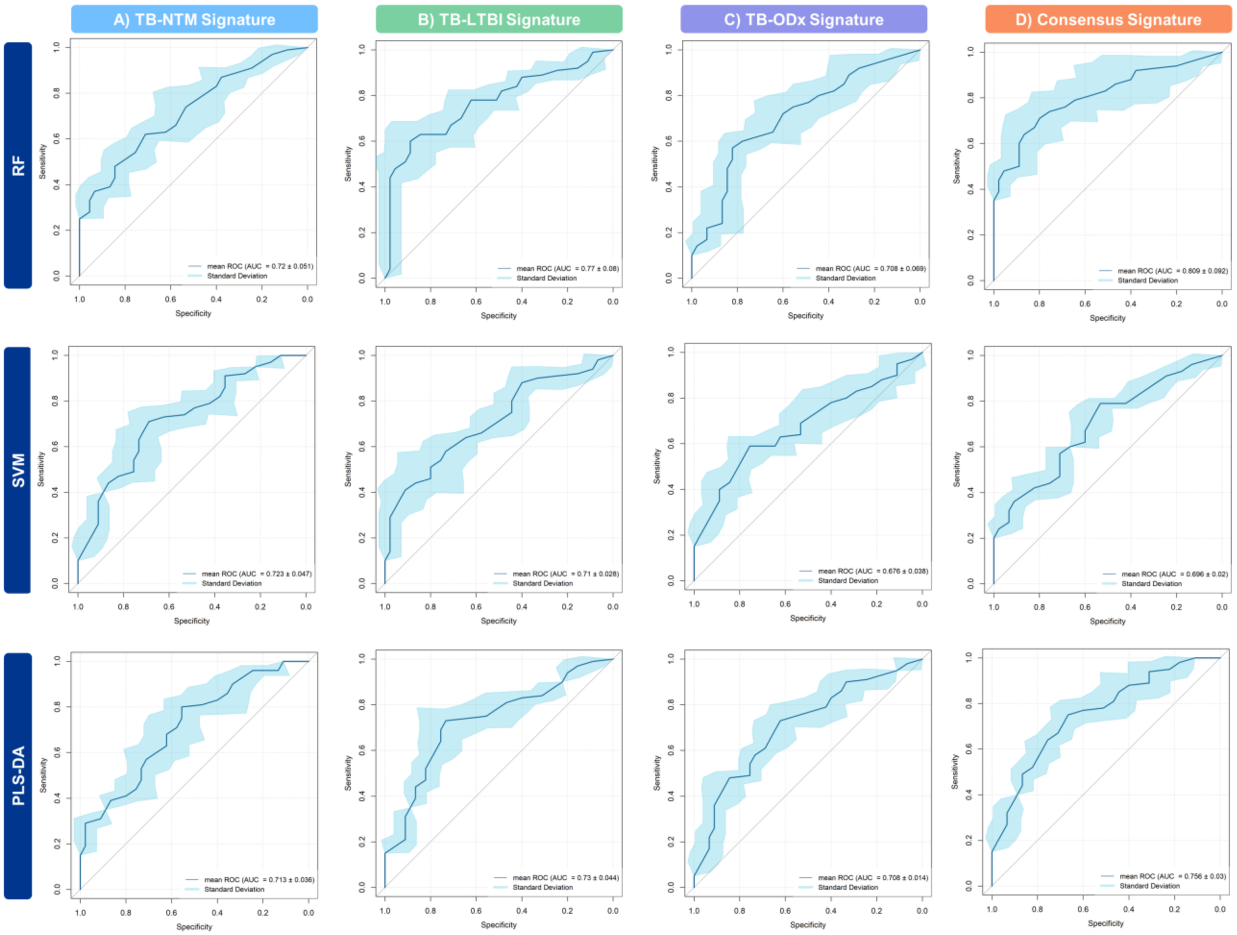

**Figure S13. Receiver operating characteristic curve of the random forest (RF), support vector machines (SVM), and partial least squares discriminant analysis (PLS-DA) models using the TB-LTBI biosignature or the TB-ODx biosignature, without clinical covariates, for the classification of TB and NTM.** Abbreviations: TB: tuberculosis; NTM: nontuberculous mycobacteria infection; LTBI: latent tuberculosis infection; and ODx: other lung diseases.

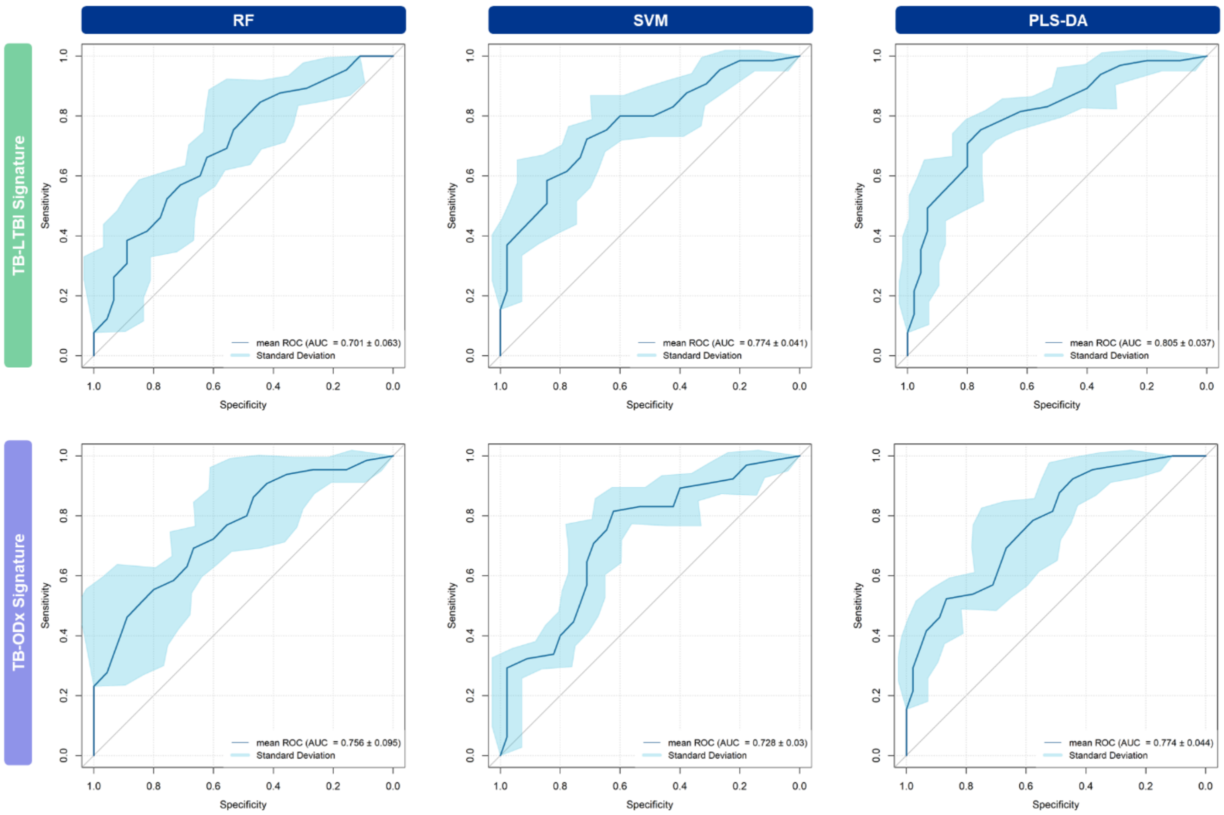

**Figure S14. Receiver operating characteristic curve of the random forest (RF), support vector machines (SVM), and partial least squares discriminant analysis (PLS-DA) models using the TB-NTM biosignature or the TB-ODx biosignature, without clinical covariates, for the classification of TB and LTBI.** Abbreviations: TB: tuberculosis; NTM: nontuberculous mycobacteria infection; LTBI: latent tuberculosis infection; and ODx: other lung diseases.

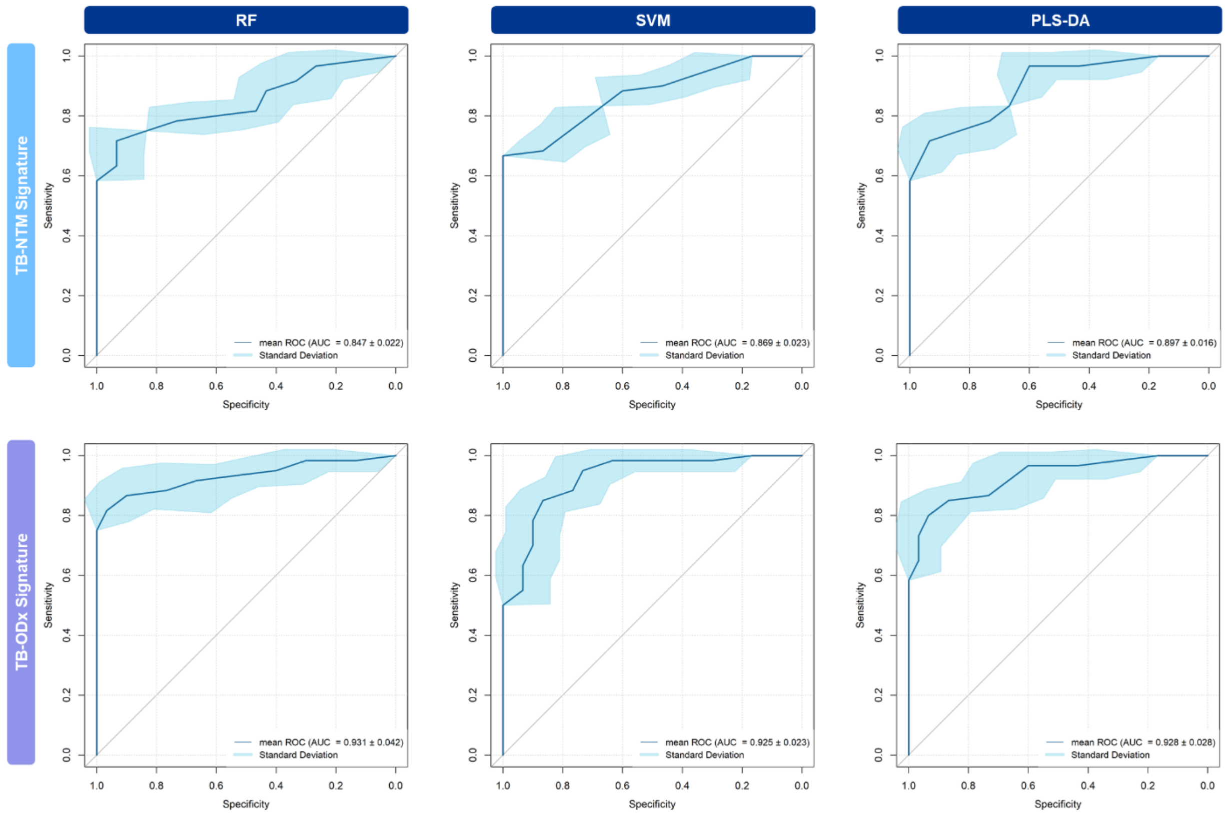

**Figure S15. Receiver operating characteristic curve of the random forest (RF), support vector machines (SVM), and partial least squares discriminant analysis (PLS-DA) models using the TB-LTBI biosignature and the TB-ODx biosignature, without clinical covariates, for the classification of TB and ODx.** Abbreviations: TB: tuberculosis; NTM: nontuberculous mycobacteria infection; LTBI: latent tuberculosis infection; and ODx: other lung diseases.

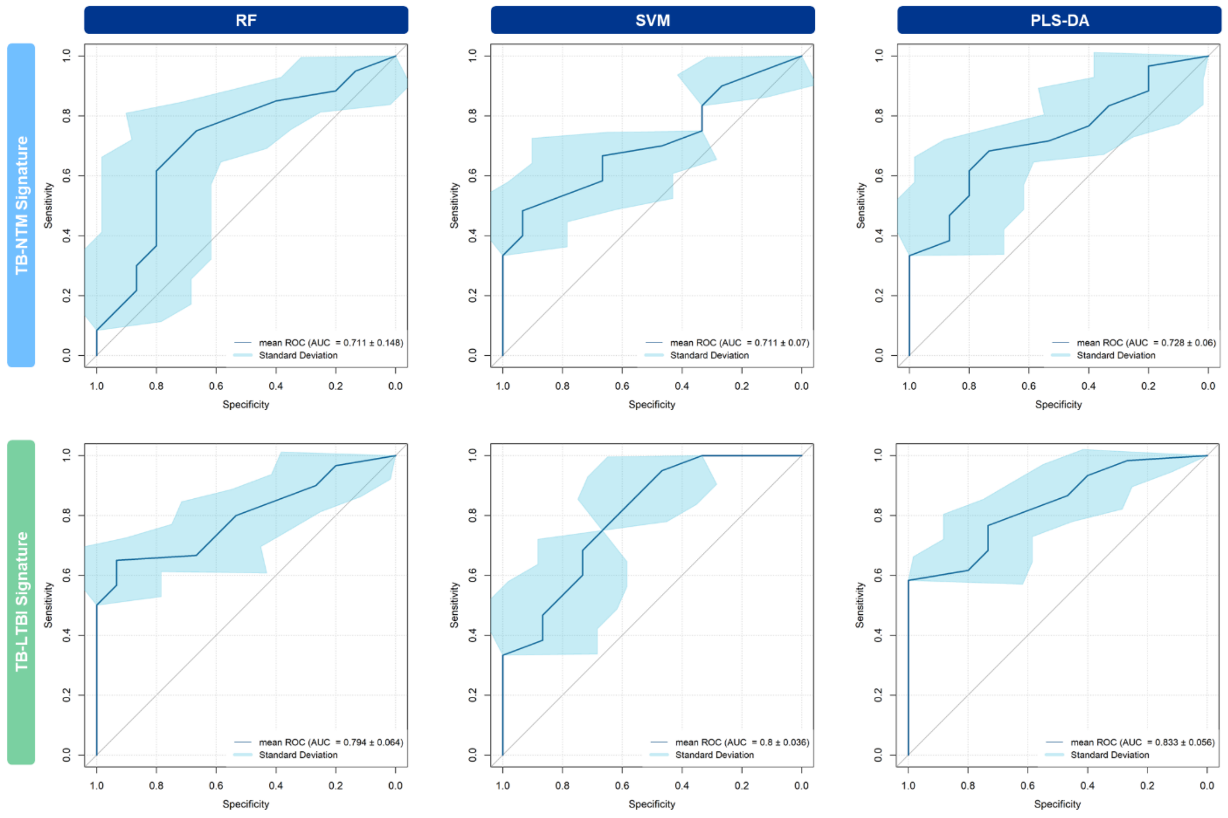

**Figure S16. Receiver operating characteristic curve of the random forest (RF), support vector machines (SVM), and partial least squares discriminant analysis (PLS-DA) models using the TB-NTM biosignature, the TB-LTBI biosignature, the TB-ODx biosignature, or the consensus biosignature, without clinical covariates, for the differentiation of TB and LTBI in the validation cohort.** Abbreviations: TB: tuberculosis; NTM: nontuberculous mycobacteria infection; LTBI: latent tuberculosis infection; and ODx: other lung diseases.

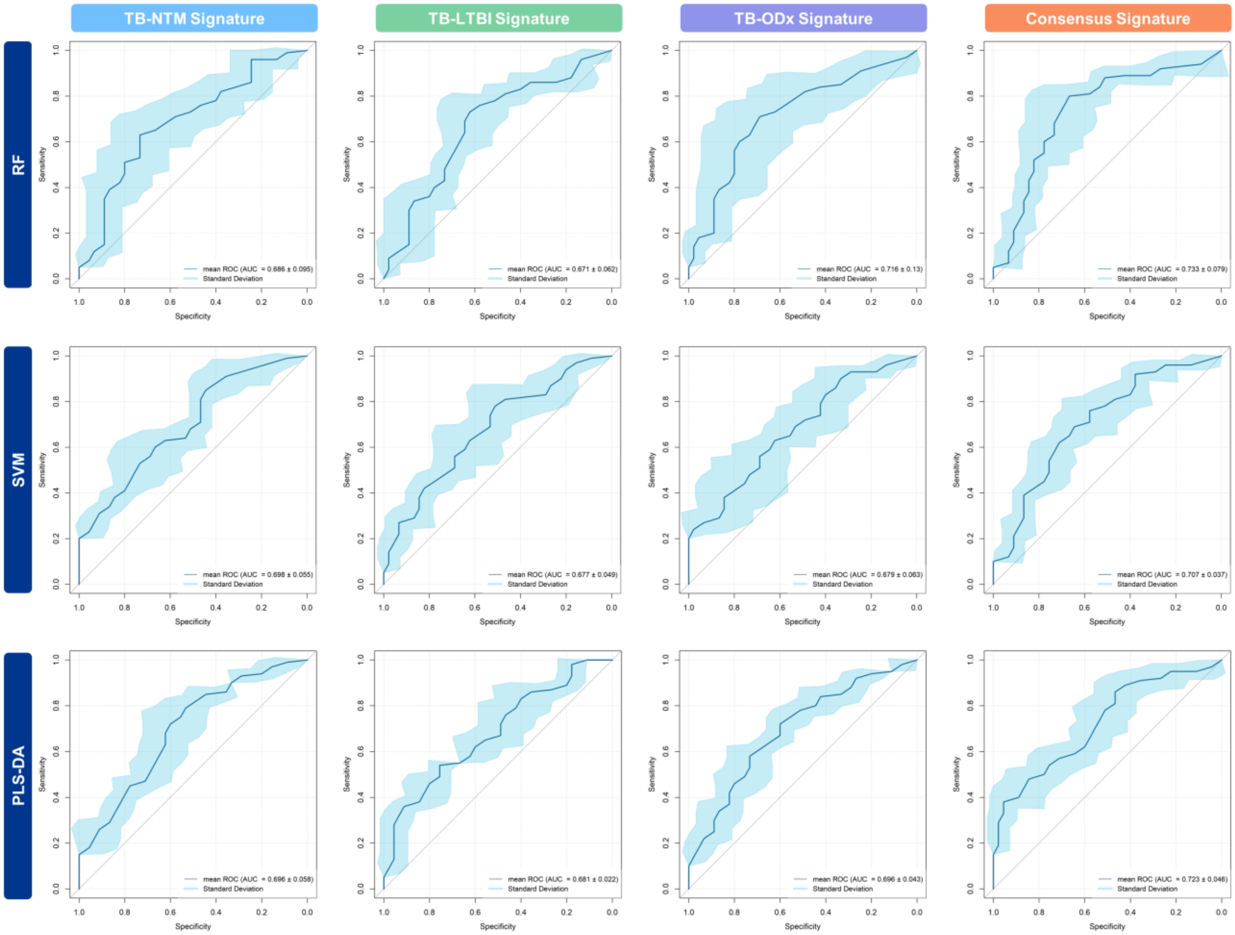

**Figure S17. Top 10% variables based on the importance score of three machine learning models of the random forest (RF), support vector machines (SVM), and partial least squares discriminant analysis (PLS-DA) for the classification of TB versus non-TB controls (NTM, LTBI, and ODx) using the TB-NTM biosignature, TB-LTBI biosignature, or TB-ODx biosignature, without clinical covariates.** Abbreviations: TB: tuberculosis; NTM: nontuberculous mycobacteria infection; LTBI: latent tuberculosis infection; and ODx: other lung diseases.

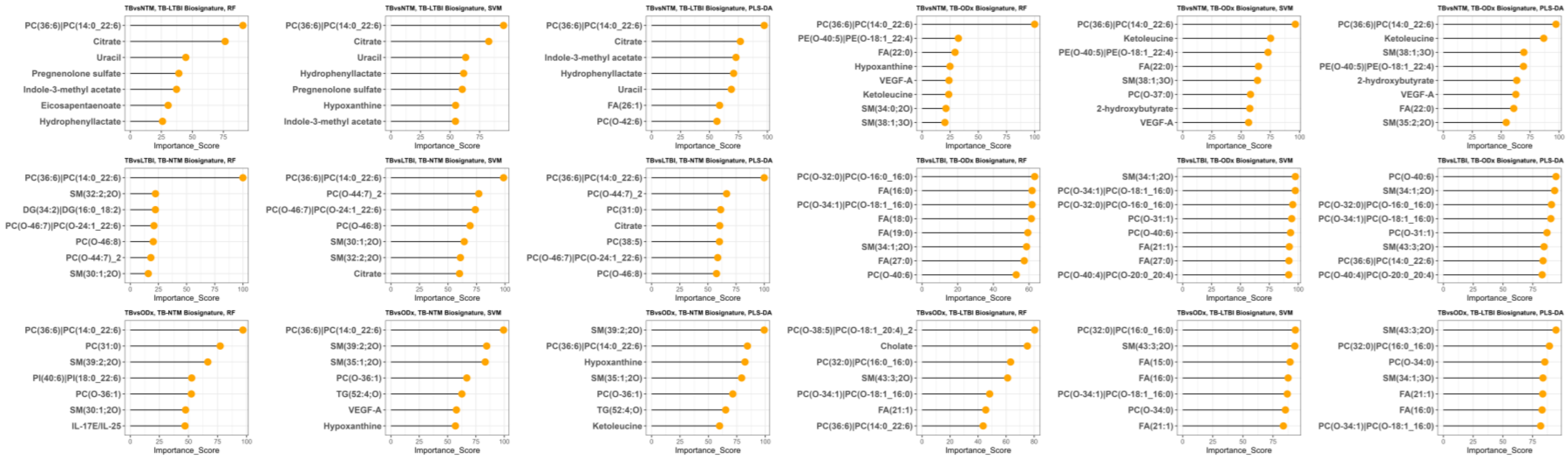

### SUPPLEMENTARY TABLES

**Table S1. Clinical characteristics of the validation cohort.**

| | Validation Cohort | | |
|---|---|---|---|
| | **All (N = 98)** | **TB group (N = 68)** | **LTBI group (N = 30)** |
| **Age, years** | 60.00 (51.25, 65.75) | 59.00 (51.00, 68.00) | 61.50 (52.75, 64.00) |
| **BMI, $kg.m^{-2}$** | 23.15 (20.93, 25.18) | 22.40 (20.50, 24.00) | 24.50 (23.55, 26.08)*** |
| **Sex** | | | |
| Male | 69 (70.41%) | 56 (82.35%) | 13 (43.33%)*** |
| Female | 29 (29.59%) | 12 (17.65%) | 17 (56.67%)*** |
| **Comorbidity** | | | |
| Diabetes | 26 (26.53%) | 21 (30.88%) | 5 (16.67%) |
| Hypertension | 51 (52.04%) | 30 (44.12%) | 21 (70.00%)* |
| **Smoking Status** | | | |
| Never | 34 (34.69%) | 21 (30.88%) | 13 (43.33%)* |
| Current | 35 (35.71%) | 31 (45.59%) | 4 (13.33%)* |
| Former | 21 (21.43%) | 15 (22.06%) | 6 (20.00%)* |
| Unknown | 8 (8.16%) | 1 (1.47%) | 7 (23.33%)* |
| **Alcohol consumption** | | | |
| Never | 24 (24.49%) | 18 (26.47%) | 6 (20.00%)* |
| Social | 19 (19.39%) | 19 (27.94%) | 0 (0.00%)* |
| Heavy | 18 (18.37%) | 16 (23.53%) | 2 (6.67%)* |
| Unknown | 37 (37.76%) | 15 (22.06%) | 22 (73.33%)* |

Abbreviation: LTBI: Latent tuberculosis infection; TB: Tuberculosis.
* P-value < 0.05, ** P-value < 0.01, *** P-value < 0.001. Data are presented as n (%) or median (interquartile range). Cohort characteristics were compared using Kruskal–Wallis rank-sum test for continuous variables and Fisher's exact test for categorical variables. Unknown values of variables were removed before conducting the statistical tests.

**Table S2. Performance of multi-ome biosignatures without clinical covariates for the classification of active TB and non-TB.**

| Biosignature | AUC ± SD | | | | | | | | |
|---|---|---|---|---|---|---|---|---|---|
| | TB vs NTM Classification | | | TB vs LTBI Classification | | | TB vs ODx Classification | | |
| | RF | SVM | PLS-DA | RF | SVM | PLS-DA | RF | SVM | PLS-DA |
| **TB-NTM Biosignature** | | | | 0.85 ± 0.02 | 0.87 ± 0.02 | 0.90 ± 0.02 | 0.71 ± 0.15 | 0.71 ± 0.07 | 0.73 ± 0.06 |
| **TB-LTBI Biosignature** | 0.70 ± 0.06 | 0.77 ± 0.04 | 0.81 ± 0.04 | | | | 0.79 ± 0.06 | 0.80 ± 0.04 | 0.83 ± 0.06 |
| **TB-ODx Biosignature** | 0.76 ± 0.10 | 0.73 ± 0.03 | 0.78 ± 0.04 | 0.93 ± 0.04 | 0.93 ± 0.02 | 0.93 ± 0.03 | | | |

Abbreviation: AUC: Area under the receiver operating characteristic curve; LTBI: Latent tuberculosis infection; NTM: Nontuberculous mycobacteria; PLS-DA: Partial least squares discriminant analysis; ODx: Other diseases; TB: Tuberculosis; RF: Random forest; SD: Standard deviation; SVM: Support vector machines.

**Table S3. Performance of multi-ome biosignatures without clinical covariates for the differentiation of TB and LTBI in the validation cohort.**

| Model | AUC ± SD | | | |
|---|---|---|---|---|
| | TB-NTM Biosignature | TB-LTBI Biosignature | TB-ODx Biosignature | Consensus Biosignature |
| **RF** | 0.69 ± 0.1 | 0.67 ± 0.06 | 0.72 ± 0.1 | 0.73 ± 0.08 |
| **SVM** | 0.70 ± 0.06 | 0.68 ± 0.05 | 0.68 ± 0.06 | 0.71 ± 0.04 |
| **PLS-DA** | 0.70 ± 0.06 | 0.68 ± 0.02 | 0.70 ± 0.04 | 0.72 ± 0.05 |

Abbreviation: AUC: Area under the receiver operating characteristic curve; LTBI: Latent tuberculosis infection; NTM: Nontuberculous mycobacteria; PLS-DA: Partial least squares discriminant analysis; ODx: Other diseases; TB: Tuberculosis; RF: Random forest; SD: Standard deviation; SVM: Support vector machines.

**Table S4. Machine learning important score-derived median rank and relative frequency of biomarkers observed in 18 scenarios of the discovery cohorts.**

| Biomarker | With clinical covariates | | Without clinical covariates | |
|---|---|---|---|---|
| | Median Ranking | Relative Frequency | Median Ranking | Relative Frequency |
| PC(14:0_22:6) | 3 | 18/18 | 1 | 18/18 |
| SM(35:1;2O) | 6 | 6/18 | 9 | 6/18 |
| PC(O-44:7)_2 | 6.5 | 6/18 | 8 | 6/18 |
| PC(31:0) | 9.5 | 6/18 | 9.5 | 6/18 |
| PC(O-46:8) | 9.5 | 6/18 | 10 | 6/18 |
| PC(O-24:1_22:6) | 10.5 | 6/18 | 11.5 | 6/18 |
| FA(21:1) | 11 | 12/18 | 9 | 12/18 |
| FA(22:0) | 11 | 6/18 | 12.5 | 6/18 |
| PI(18:0_22:6) | 13.5 | 6/18 | 11 | 6/18 |
| SM(39:1;2O) | 14 | 6/18 | 14 | 6/18 |
| SM(30:1;2O) | 14.5 | 6/18 | 10 | 6/18 |
| Citrate | 15.5 | 12/18 | 28 | 12/18 |
| FA(20:0) | 15.5 | 12/18 | 18.5 | 12/18 |
| FA(26:1) | 15.5 | 6/18 | 10 | 6/18 |
| SM(43:3;2O) | 16 | 12/18 | 11.5 | 12/18 |
| PC(O-36:1) | 16.5 | 6/18 | 14 | 6/18 |
| SM(39:2;2O) | 16.5 | 6/18 | 12 | 6/18 |
| FA(27:0) | 17.5 | 6/18 | 18.5 | 6/18 |
| FA(18:0) | 18.5 | 12/18 | 23 | 12/18 |
| Hydrophenyllactate | 19 | 12/18 | 12.5 | 12/18 |
| Hypoxanthine | 19 | 18/18 | 16.5 | 18/18 |
| PC(38:5) | 19.5 | 6/18 | 21.5 | 6/18 |
| PE(O-18:1_22:4) | 21 | 6/18 | 18 | 6/18 |
| FA(19:0) | 21.5 | 12/18 | 19.5 | 12/18 |
| DG(16:0_18:2) | 22 | 6/18 | 16.5 | 6/18 |
| PC(O-40:4) | 22.5 | 6/18 | 20.5 | 6/18 |
| Cer(18:1;2O/16:0) | 23 | 12/18 | 19 | 12/18 |
| FA(15:0) | 23 | 12/18 | 26.5 | 12/18 |
| FA(16:0) | 23 | 12/18 | 19.5 | 12/18 |
| Ketoleucine | 23 | 12/18 | 21 | 12/18 |
| PC(O-18:1_16:0) | 24.5 | 12/18 | 19.5 | 12/18 |
| Indole-3-methyl acetate | 25 | 12/18 | 46.5 | 12/18 |
| PC(O-42:6) | 25 | 6/18 | 21.5 | 6/18 |
| SM(32:2;2O) | 25 | 6/18 | 35.5 | 6/18 |
| CAR(8:1) | 25.5 | 6/18 | 31 | 6/18 |
| Choline | 25.5 | 6/18 | 33.5 | 6/18 |
| PC(O-40:6) | 25.5 | 12/18 | 23 | 12/18 |
| DG(16:0_18:1) | 26.5 | 6/18 | 24.5 | 6/18 |

| | | | | |
|---|---|---|---|---|
| Uracil | 26.5 | 12/18 | 25 | 12/18 |
| SM(34:0;2O) | 27 | 6/18 | 19.5 | 6/18 |
| PC(O-35:0) | 28 | 6/18 | 35 | 6/18 |
| PC(O-34:0) | 28.5 | 12/18 | 20.5 | 12/18 |
| Pregnenolone sulfate | 29 | 6/18 | 37 | 6/18 |
| Eicosapentaenoate | 30 | 12/18 | 18 | 12/18 |
| PC(O-16:0_16:0) | 30 | 12/18 | 26.5 | 12/18 |
| Hippurate | 30.5 | 6/18 | 36.5 | 6/18 |
| SM(40:3;2O) | 31 | 6/18 | 30.5 | 6/18 |
| PC(16:0_22:6) | 31.5 | 6/18 | 28.5 | 6/18 |
| PC(O-32:1)_2 | 31.5 | 6/18 | 34 | 6/18 |
| Deoxycholate | 32 | 12/18 | 33 | 12/18 |
| PC(O-20:0_20:4) | 32 | 12/18 | 28 | 12/18 |
| Gende: Female | 33 | 18/18 | - | - |
| Gende: Male | 33 | 18/18 | - | - |
| SM(35:2;2O) | 33 | 6/18 | 30.5 | 6/18 |
| SM(37:6;2O) | 33 | 12/18 | 40 | 12/18 |
| LPE(20:5) | 33.5 | 12/18 | 40 | 12/18 |
| PC(O-44:6)_2 | 33.5 | 6/18 | 28.5 | 6/18 |
| Hex3Cer(18:1;2O/16:0) | 34 | 6/18 | 29 | 6/18 |
| O-acetylserine | 34 | 6/18 | 20.5 | 6/18 |
| PC(O-37:0) | 34 | 6/18 | 38.5 | 6/18 |
| PC(O-40:7) | 34.5 | 6/18 | 36 | 6/18 |
| Indole-3-carboxyaldehyde | 35 | 12/18 | 33.5 | 12/18 |
| Cer(44:1;3O) | 35.5 | 6/18 | 35.5 | 6/18 |
| TG(16:0_18:2_20:4) | 35.5 | 6/18 | 34 | 6/18 |
| PC(O-18:1_20:4)_2 | 36 | 6/18 | 39 | 6/18 |
| Age | 36.5 | 18/18 | - | - |
| CAR(10:0) | 37 | 12/18 | 47.5 | 12/18 |
| MIG | 37.5 | 12/18 | 31 | 12/18 |
| 6-hydroxydopamine | 38 | 12/18 | 40 | 12/18 |
| IL-15 | 38.5 | 18/18 | 37 | 18/18 |
| Thyroxine | 38.5 | 6/18 | 35 | 6/18 |
| BMI | 39.5 | 18/18 | - | - |
| PC(16:0_16:0) | 39.5 | 12/18 | 36 | 12/18 |
| TG(52:4;O) | 39.5 | 6/18 | 29.5 | 6/18 |
| Cholate | 40 | 12/18 | 42.5 | 12/18 |
| SM(34:1;3O) | 40 | 12/18 | 35.5 | 12/18 |
| Dihydrouracil | 40.5 | 6/18 | 46.5 | 6/18 |
| PC(O-24:1_20:4) | 40.5 | 6/18 | 42 | 6/18 |
| Valine | 40.5 | 12/18 | 41.5 | 12/18 |
| PC(16:0_18:0) | 41 | 6/18 | 32.5 | 6/18 |
| SE(29:1/20:4)_2 | 41.5 | 6/18 | 33 | 6/18 |
| PC(18:1_22:6) | 42 | 6/18 | 45.5 | 6/18 |

| | | | | |
|---|---|---|---|---|
| PC(O-44:6)_1 | 42 | 6/18 | 39.5 | 6/18 |
| Tryptophan | 42 | 12/18 | 45.5 | 12/18 |
| CAR(18:1) | 42.5 | 6/18 | 48.5 | 6/18 |
| PC(15:0_22:6) | 42.5 | 6/18 | 40 | 6/18 |
| PC(O-16:1_22:6) | 42.5 | 6/18 | 38 | 6/18 |
| PC(O-22:0_20:4) | 42.5 | 6/18 | 54.5 | 6/18 |
| 3-nitro-tyrosine | 43 | 12/18 | 40 | 12/18 |
| CAR(12:1) | 43 | 12/18 | 53 | 12/18 |
| CAR(8:0) | 43 | 6/18 | 44.5 | 6/18 |
| PC(18:0_22:5) | 43 | 6/18 | 47.5 | 6/18 |
| TG(16:0_18:1_18:2)_2 | 43 | 6/18 | 47 | 6/18 |
| TG(O-51:1) | 43 | 6/18 | 50.5 | 6/18 |
| 2-hydroxybutyrate | 43.5 | 12/18 | 45 | 12/18 |
| PC(O-40:5)\|PC(O-20:1_20:4) | 43.5 | 6/18 | 59 | 6/18 |
| CAR(14:2) | 44 | 12/18 | 38.5 | 12/18 |
| IL-1RA | 44 | 12/18 | 38 | 12/18 |
| CE(20:4) | 44.5 | 6/18 | 40 | 6/18 |
| PE(34:1)\|PE(16:0_18:1) | 44.5 | 6/18 | 45 | 6/18 |
| CAR(18:2) | 45 | 6/18 | 57 | 6/18 |
| O-succinyl-homoserine | 45 | 12/18 | 26.5 | 12/18 |
| (2-aminoethyl)phosphonate | 45.5 | 6/18 | 39 | 6/18 |
| Dehydroisoandrosterone sulfate | 45.5 | 6/18 | 58 | 6/18 |
| PC(O-22:2_20:4) | 45.5 | 6/18 | 39.5 | 6/18 |
| Lactate | 46 | 6/18 | 45 | 6/18 |
| Cer(18:1;2O/24:0) | 46.5 | 6/18 | 47 | 6/18 |
| SM(42:3;2O) | 46.5 | 6/18 | 42.5 | 6/18 |
| IP-10 | 47 | 12/18 | 44.5 | 12/18 |
| CAR(14:1) | 47.5 | 12/18 | 47 | 12/18 |
| PC(O-34:6) | 47.5 | 6/18 | 35 | 6/18 |
| PC(O-22:1_20:4) | 47.5 | 6/18 | 34 | 6/18 |
| SM(38:1;3O) | 48.5 | 6/18 | 41 | 6/18 |
| TG(16:0_18:1_20:4) | 48.5 | 6/18 | 50.5 | 6/18 |
| CAR(11:1) | 49 | 6/18 | 53.5 | 6/18 |
| Ribose 1,5-bisphosphate | 49 | 6/18 | 48.5 | 6/18 |
| SM(42:2;2O)_2 | 49 | 12/18 | 40.5 | 12/18 |
| PC(O-31:1) | 49.5 | 12/18 | 38 | 12/18 |
| SM(33:2;3O) | 49.5 | 6/18 | 40 | 6/18 |
| Taurodeoxycholate | 50.5 | 6/18 | 58 | 6/18 |
| CAR(12:0) | 51 | 6/18 | 50 | 6/18 |
| Kynurenate | 51 | 6/18 | 47 | 6/18 |
| PC(41:3) | 51.5 | 6/18 | 39 | 6/18 |
| SM(42:2;2O)_1 | 51.5 | 6/18 | 42 | 6/18 |
| Hex2Cer(18:1;2O/16:0) | 52.5 | 6/18 | 52.5 | 6/18 |
| TG(52:2) | 52.5 | 6/18 | 44.5 | 6/18 |

| | | | | |
|---|---|---|---|---|
| Cysteate | 53 | 6/18 | 66.5 | 6/18 |
| MCP-3 | 53 | 6/18 | 55 | 6/18 |
| SM(34:1;2O) | 53 | 12/18 | 34.5 | 12/18 |
| Docosahexaenoate | 53.5 | 6/18 | 45.5 | 6/18 |
| Glutamine | 53.5 | 6/18 | 53 | 6/18 |
| VEGF-A | 53.5 | 12/18 | 41 | 12/18 |
| PC(18:0_20:4) | 54 | 6/18 | 52.5 | 6/18 |
| Hexitol | 54.5 | 6/18 | 57 | 6/18 |
| CAR(3:0) | 55 | 6/18 | 60.5 | 6/18 |
| GRO alpha | 55 | 6/18 | 68.5 | 6/18 |
| IL-17E/IL-25 | 55.5 | 6/18 | 57.5 | 6/18 |
| PDGF-AB/BB | 55.5 | 6/18 | 59 | 6/18 |
| Indoxyl sulfate | 56 | 6/18 | 51 | 6/18 |
| SM(31:1;2O) | 56 | 6/18 | 43 | 6/18 |
| SM(34:2;2O) | 56.5 | 6/18 | 42 | 6/18 |
| 3-(2-hydroxyphenyl)propanoate | 57 | 6/18 | 59 | 6/18 |
| Citrulline | 57 | 6/18 | 60 | 6/18 |
| Trimethylamine N-oxide | 57 | 6/18 | 44 | 6/18 |
| Glutamate | 57.5 | 6/18 | 50.5 | 6/18 |
| IL-8 | 57.5 | 6/18 | 51 | 6/18 |
| Indolelactate | 57.5 | 6/18 | 64 | 6/18 |
| Chenodeoxycholate | 58 | 6/18 | 49.5 | 6/18 |
| CAR(5:0) | 59 | 6/18 | 73 | 6/18 |
| Eotaxin | 59.5 | 6/18 | 60 | 6/18 |
| LPE(O-18:2) | 59.5 | 6/18 | 63 | 6/18 |
| SM(33:1;3O) | 59.5 | 12/18 | 48 | 12/18 |
| Hexose | 60 | 6/18 | 53.5 | 6/18 |
| IFN gamma | 60 | 3/18 | 60 | 3/18 |
| PC(39:5) | 60 | 6/18 | 63 | 6/18 |
| Cer(18:1;2O/22:0) | 60.5 | 6/18 | 50.5 | 6/18 |
| LPE(O-16:1) | 60.5 | 6/18 | 55.5 | 6/18 |
| PC(33:2;3O) | 60.5 | 6/18 | 49.5 | 6/18 |
| Theophylline | 60.5 | 12/18 | 50 | 12/18 |
| IL-9 | 61 | 12/18 | 51 | 12/18 |
| Phenylacetylglutamine | 62 | 6/18 | 57.5 | 6/18 |
| PC(18:0_22:6) | 62.5 | 6/18 | 67 | 6/18 |
| EGF | 63 | 6/18 | 64.5 | 6/18 |
| IFN alpha2 | 64 | 6/18 | 54.5 | 6/18 |
| TG(18:1_18:1_20:4) | 64 | 6/18 | 72.5 | 6/18 |
| Histidine | 64.5 | 12/18 | 65 | 12/18 |
| SM(36:1;2O) | 64.5 | 6/18 | 44.5 | 6/18 |
| Deoxyadenosine monophosphate | 65 | 6/18 | 71.5 | 6/18 |
| TG(16:0_18:0_18:1)_2 | 65 | 6/18 | 64.5 | 6/18 |

| | | | | |
|---|---|---|---|---|
| TG(16:0_18:1_18:1)_2 | 65.5 | 6/18 | 55.5 | 6/18 |
| IL-3 | 66 | 6/18 | 65.5 | 6/18 |
| Ornithine | 66.5 | 6/18 | 61 | 6/18 |
| TNF alpha | 67 | 6/18 | 54 | 6/18 |
| Taurine | 67 | 6/18 | 67.5 | 6/18 |
| FGF-2 | 69 | 6/18 | 56.5 | 6/18 |
| Fractalkine | 69 | 6/18 | 62 | 6/18 |
| CAR(16:0) | 70 | 6/18 | 64.5 | 6/18 |
| TG(O-17:0_18:1_20:4) | 70.5 | 6/18 | 52.5 | 6/18 |
| IL-18 | 71 | 6/18 | 63.5 | 6/18 |
| Betaine | 71.5 | 6/18 | 50 | 6/18 |
| Indole-3-acetate | 71.5 | 6/18 | 62.5 | 6/18 |
| Urea | 72 | 6/18 | 73.5 | 6/18 |
| Glycochenodeoxycholate | 72.5 | 6/18 | 70 | 6/18 |
| MIP-1 alpha | 74 | 6/18 | 60 | 6/18 |
| Taurocholate | 74.5 | 6/18 | 69.5 | 6/18 |
| IL-1 beta | 75 | 6/18 | 69 | 6/18 |
| TG(O-19:1_18:0_18:0) | 77.5 | 6/18 | 62.5 | 6/18 |
